# A Preoperative Electroencephalography Signature for Predicting Treatment Response to Deep Brain Stimulation in Obsessive-Compulsive Disorder

**DOI:** 10.64898/2026.03.03.26347351

**Authors:** Wenlong Wang, Jiayue Cheng, Xin Zhang, Xi Wu, Hanyang Ruan, Binxin Huang, Tingting Xu, Feifei Qi, Yan Liang, Hongyi Zhi, Jian Gao, Luolong Cao, Yang Wang, Kaiming Zhuo, Corey J. Keller, Gerwin Schalk, Jing Jiang, Qing Fan, Nolan Williams, Huiqin Han, Wei Wu, Zhen Wang

**Author notes:** These authors contributed equally (Wenlong Wang, Jiayue Cheng, Xin Zhang, Xi Wu).

## Abstract

Deep brain stimulation (DBS) is effective for treatment-refractory obsessive-compulsive disorder (OCD), but outcomes are heterogeneous and non-responders incur surgical and financial burden. We sought a scalable, non-invasive preoperative signature of DBS response. Using preoperative resting-state electroencephalography (EEG) from a randomized, double-blind, sham-controlled trial of nucleus accumbens/anterior limb of internal capsule DBS (*N* = 24; NCT04967560), we developed a machine learning approach tailored to small samples to identify lower relative delta power at a right fronto-temporal electrode as a robust predictor of greater six-month symptom reduction, explaining >40% of outcome variance and improving the response rate >20% over all-comers. This EEG signature was aberrant relative to healthy controls yet unrelated to baseline symptom severity, and predictive only under active, not sham, stimulation. Notably, it prospectively predicted outcomes in an independent cohort (7 of 8 patients). The signature generalized across eyes-open and eye-closed recordings, was corroborated by source-space magnetoencephalography, and showed excellent short- and long-term test-retest reliability. Mechanistically, this EEG signature was highly correlated with right fronto-temporal aperiodic exponent, and its predictive strength was concentrated in cortical regions enriched for inhibitory-neuron markers, linking the signature to excitation-inhibition balance. Moreover, longitudinal signature changes tracked clinical improvement, providing proof-of-concept support for its use as a target-engagement readout. These findings establish a scalable, biologically grounded EEG signature to guide patient selection for DBS in severe OCD.

**One-sentence summary:** Preoperative resting-state EEG delta power predicts which OCD patients respond to deep brain stimulation.

**Trial Registration:** The Efficacy and Safety of ALIC/NAcc-DBS for Treatment-Refractory OCD, NCT04967560

## Introduction

Obsessive-Compulsive Disorder (OCD) is a chronic and often debilitating psychiatric condition characterized by intrusive thoughts and repetitive, compulsive behaviors that can severely impair an individual’s quality of life, with a lifetime prevalence of 2.3%^1^. While first-line treatments such as serotonin reuptake inhibitors (SSRIs) and cognitive-behavioral therapy (CBT) are effective for many, a significant portion of patients remain treatment-refractory^2^. For these individuals, deep brain stimulation (DBS) has emerged as a powerful therapeutic option^3–5^.

By targeting key nodes of the cortico-striato-thalamo-cortical (CSTC) circuits^6^, such as the anterior limb of the internal capsule (ALIC), subthalamic nucleus (STN), nucleus accumbens (NAcc), and ventral capsule/ventral striatum (VC/VS), DBS has shown considerable efficacy across multiple clinical trials, with response rates reaching 50-70%^7^. Early studies demonstrated success targeting either the ALIC, a major white matter pathway, or the NAcc, a critical gray matter structure for reward and motivation^8,9^. More recently, a therapeutic strategy of stimulating both the NAcc and ALIC in conjunction has gained traction^10^, based on the rationale that modulating both gray matter function and white matter connectivity may yield superior clinical outcomes. Indeed, recent trials investigating this combined target have reported promising results, further establishing DBS as a viable intervention for this difficult-to-treat population^11^.

However, the application of DBS presents significant challenges. The procedure is surgically invasive with inherent risks and represents a substantial financial investment for both patients and healthcare systems. Compounding this is the notable variability in clinical outcomes, with not all patients experiencing a significant reduction in symptoms^12^. This underscores a critical and unmet need in the field: the development of non-invasive, reliable, preoperative signature to predict therapeutic success^13^. An accurate treatment-predictive signature would substantially enhance patient care by enabling clinicians to identify which individuals are most likely to benefit from DBS before they undergo the costly and invasive procedure. This would not only maximize the therapeutic potential of DBS by directing it toward the most suitable candidates but also spare non-responders from an unnecessary intervention.

To date, the most compelling treatment-predictive findings have emerged from connectomic studies, which leverage both anatomic and functional neuroimaging methods to identify disease-relevant neural circuits^14^. Diffusion magnetic resonance imaging (dMRI) has been instrumental in this effort by enabling the mapping of white-matter pathways. Using this anatomical approach, studies have found that connectivity to specific cortical regions can predict clinical outcomes^15–18^. For instance, a key study showed that connectivity to the medial and lateral prefrontal cortices and a specific frontothalamic pathway predicted clinical improvement in DBS for OCD^16^. Another study identified a common fiber bundle that predicted clinical improvement across different DBS targets, including ALIC, NAcc, and STN^15^ (However, this view is complicated by reports that VC and STN stimulation can differentially impact mood versus cognitive flexibility^19^). A large-scale, transdiagnostic study mapped dysfunctional circuits in the frontal cortex and found a topographical arrangement of connections, with OCD specifically involving circuits with the ventromedial prefrontal and anterior cingulate cortices^17^. Additionally, functional connectomics using functional MRI (fMRI) has demonstrated that DBS can induce a reduction in functional connectivity between the NAcc and prefrontal cortices, an effect that correlates with improved outcomes^18^. Recently, intracranial recordings from implanted electrodes have identified a treatment-predictive signature based on the disruption of neural periodicity in the VS^20^. While powerful, such intracranial signatures are inherently invasive and can only be measured post-operatively, limiting their clinical utility for pre-surgical patient selection.

EEG represents a particularly promising yet under-explored modality for DBS treatment prediction in OCD. As a non-invasive, cost-effective, and widely accessible technology, EEG offers a highly feasible screening tool. Its excellent temporal resolution allows for the detailed capture of brain dynamics that may predict a patient’s capacity to respond to neuromodulation^21^. A leading neurobiological theory posits that OCD arises from a cortical excitation/inhibition (E/I) imbalance, particularly within frontal circuits, leading to a state of hyperexcitability^22^. Several EEG-based measures are thought to be electrophysiological signatures of this imbalance. For example, the Error-Related Negativity (ERN), an EEG signal reflecting hyperactive performance monitoring, is consistently exaggerated in OCD and has been shown to predict response to SSRIs and CBT^23^. Other measures, such as resting-state frontal alpha asymmetry and power in various frequency bands, have also been explored as potential predictors, possibly reflecting deficits in cortical inhibition^21,24^. Importantly, changes in prefrontal delta band (1-4 Hz) oscillations in response to active DBS applied to the VC/VS have been shown to correlate with treatment success, suggesting that EEG power in this low-frequency range may be a valuable indicator of therapeutic engagement with disease-relevant circuits^25^.In addition to these oscillatory and event-related measures, the aperiodic component of the EEG power spectrum may provide complementary information about broadband cortical excitability, with a flatter spectral slope often interpreted as reflecting relatively greater excitation over inhibition^26^. While these findings establish proof-of-concept for EEG-based treatment prediction, a validated preoperative EEG signature that specifically predicts response to DBS for OCD remains elusive-a critical gap given neuromodulation’s distinct mechanisms and patient population.

To achieve clinical utility, a treatment-predictive signature must also demonstrate treatment specificity^27,28^. That is, it should predict outcomes specifically for the active intervention and not for a placebo or sham condition. A measure that correlates with outcomes in both the active DBS stimulation and sham DBS stimulation groups may simply be a general prognostic factor (e.g., reflecting overall disease severity or resilience) rather than a true predictor of response to the specific mechanism of the therapy. Therefore, establishing that a signature predicts response differentially—correlating with improvement only in the active DBS group—is essential to confirm that it is linked to the neurophysiological effects of the stimulation itself. This requirement necessitates sham-controlled designs—rare in DBS research due to ethical considerations but essential for validation.

Furthermore, an ideal treatment-predictive signature should not only predict the final outcome but also track the therapeutic response over time. A signature that changes in concert with clinical improvement could serve as an objective physiological readout of target engagement^29^. Such a readout would be invaluable for optimizing stimulation parameters and is a critical prerequisite for the development of adaptive closed-loop DBS systems that can adjust stimulation in real-time based on a patient’s neural state30.

The present study reports the results from a double-blind, randomized, sham-controlled clinical trial investigating the efficacy of combined NAcc and ALIC DBS for treatment-refractory OCD (ClinicalTrials.gov registration: NCT04967560)^31^. This rigorously designed trial provides a unique and valuable discovery dataset (cohort 1) for studying treatment prediction from a cohort of 24 patients. The study began with a 3-month double-blind phase where patients were randomized to either active or sham stimulation. Following this, the trial entered a 3-month open-label phase where patients in the sham arm crossed over to receive three months of active stimulation, while those in the active arm continued to receive three months of active simulation (Fig. S1). Participants were followed longitudinally with clinical assessments and resting-state EEG, while preoperative baseline measures also included magnetoencephalography (MEG), and structural MRI. We hypothesized that the preoperative, baseline state of EEG spectral power could serve as a non-invasive predictor of a patient’s capacity to respond to neuromodulation of these circuits. To rigorously test the generalizability of the identified signature, a second, independent cohort of 8 patients (cohorts 2&3) was prospectively recruited for out-of-sample validation. Leveraging this two-stage design, the present work was carried out to systematically address the critical gaps in EEG biomarker development for OCD. Specifically, we sought to answer the following key questions:

1. Can we identify a preoperative, resting-state EEG signature that predicts clinical response to combined NAcc and ALIC DBS?
2. Does this signature demonstrate treatment specificity by predicting outcomes differentially across active DBS and sham stimulation conditions?
3. Is the identified signature stable across time points, demonstrating good test-retest reliability?
4. Is the identified signature robust and consistent across different resting-state conditions (eyes-open vs. eyes-closed) and neuroimaging modalities (EEG vs. MEG)?
5. Can the predictive utility of this signature be prospectively replicated in an independent sample of patients?
6. What is the potential neurophysiological and molecular mechanism underlying this EEG predictive signature?
7. Can DBS treatment move this signature in the desired direction, supporting its potential utility as a direct physiological readout for target engagement?

By addressing these questions systematically, this study aims to establish a prospectively validated treatment-predictive EEG signature for DBS response in OCD—a critical step toward precision neuromodulation and improved outcomes for treatment-refractory patients. The datasets, analysis tasks, and overall workflow are summarized in Fig. 1.

**Fig. 1.**
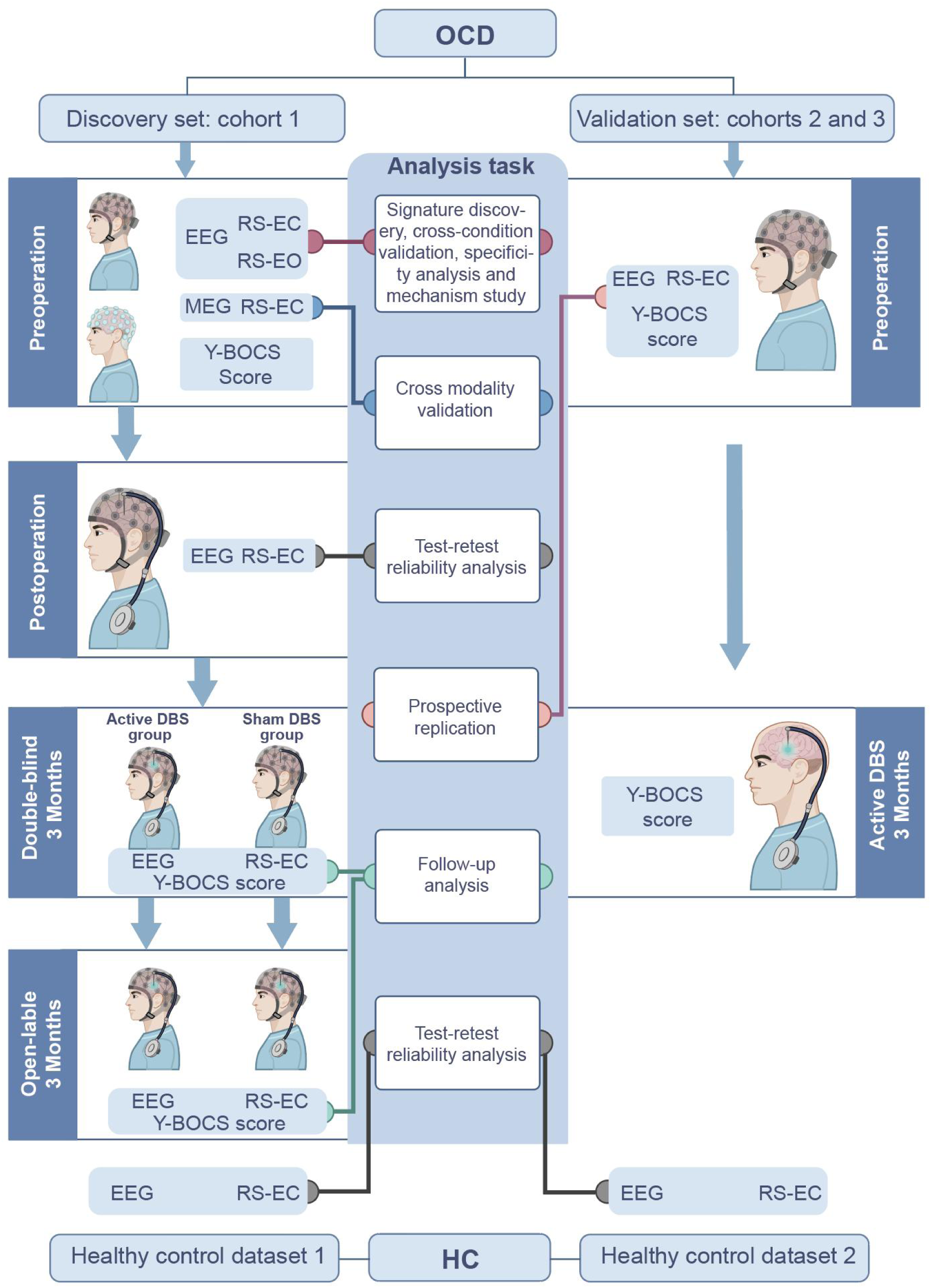
Study design and analysis workflow. Discovery data were obtained from a randomized, sham-controlled DBS trial in treatment-refractory OCD (cohort 1, *N* =24). Multimodal baseline measures (EEG, MEG) and longitudinal follow-up with resting-state EEG and Y-BOCS assessments enabled biomarker discovery, cross-condition and cross-modality validation. In addition, two healthy control (HC) datasets were included to assess short-term reliability (healthy control dataset 1, *N* =40) and long-term reliability (healthy control dataset 2, *N* =208). An independent validation set (cohorts 2 and 3, *N* =8) was used for prospective validation. RS = resting-state, EO = eyes-open, EC= eyes-closed.

## Results

### Active DBS reduces obsessive-compulsive symptoms relative sham stimulation

In the double-blind, randomized, sham-controlled clinical trial (Fig. S1), 24 patients (i.e., cohort 1) who met the criteria were recruited and randomly assigned to the active DBS group (receiving therapeutic stimulation; *N* = 12) or the sham DBS group (implanted but stimulation kept off; *N* = 12). One participant dropped out after the double-blind phase due to withdrawal of consent. The baseline characteristics of the two groups were similar (Table 1).

**Table 1.** Baseline participants demographics and characteristics. Data are n (%), mean (SD), or median (IQR). Y-BOCS = Yale-Brown Obsessive-Compulsive Scale, CGI-S = CGI-Severity, HAMD = Hamilton Depression Scale, HAMA = Hamilton Anxiety Scale, DBS = deep brain stimulation. SD = standard deviation; IQR = interquartile range.

|  | Active DBS<br>group (*N* =<br>12) | Sham DBS<br>group (*N* =<br>12) | Total (*N* = 24) |
| --- | --- | --- | --- |
| Age | 34.4 (14.0) | 29.9 (9.1) | 32.2 (11.8) |
| Sex |  |  |  |
| Male | 9 (75%) | 10 (80%) | 42 (79.2%) |
| Female | 3 (25%) | 2 (20%) | 18 (20.8%) |
| Education (year) | 16.0 (9.0–<br>16.0) | 14.0 (9.8–<br>16.0) | 16.0 (9.0–<br>16.0) |
| Course of illness | 15.8 (10.7) | 10.3 (6.3) | 13.1 (9.0) |
| BMI | 24.0 (3.5) | 24.3 (3.7) | 24.2 (3.5) |
| Y-BOCS score |  |  |  |
| Total | 29.7 (3.2) | 30.3 (3.1) | 30.0 (3.1) |
| Obsession | 14.7 (1.5) | 15.3 (1.4) | 15.0 (1.5) |
| Compulsion | 15.0 (1.8) | 15.0 (1.9) | 15.0 (1.8) |
| CGI-S | 6.0 (5.0–6.75) | 6.0 (6.0–<br>7.0) | 6.0 (5.25–7.0) |
| HAMD score | 10.33 (2.5) | 17.9 (7.2) | 14.0 (6.5) |
| HAMA score | 7.6 (5.6) | 12.2 (5.9) | 9.9 (6.1) |

At the 3-month follow-up, 5 participants in the active DBS group (41.7%) and no participant in the sham DBS group responded to treatment, indicating a significant difference (Fisher’s exact test, *p =* 0.037). The mean Y-BOCS score decreased by 8.58 points (SD 3.86, reduction rate 29.4%) in the active DBS group and by 1.00 points (SD 6.00, reduction rate 3.08%) in the sham DBS group. Mixed linear model showed a significant difference between the groups (95% CI: 3.55 to 11.62, Cohen’s d = 2.128, *p =* 0.001, Fig. 2a). At the 6-month follow-up, after the sham stimulation group had received three months of active stimulation, 62.5% (15/24) of the participants responded. There was no longer a significant difference in the mean Y-BOCS score reduction between the two groups. For CGI-S, the change of score at 3-month follow-up between active DBS and sham DBS groups was also significance (95% CI: 0.07 to 1.43, Cohen’s d = 1.251, *p* = 0.036, Fig. 2d). For HAMA and HAMD, no significant differences were observed (Fig. 2b and 2c).

**Fig. 2.**
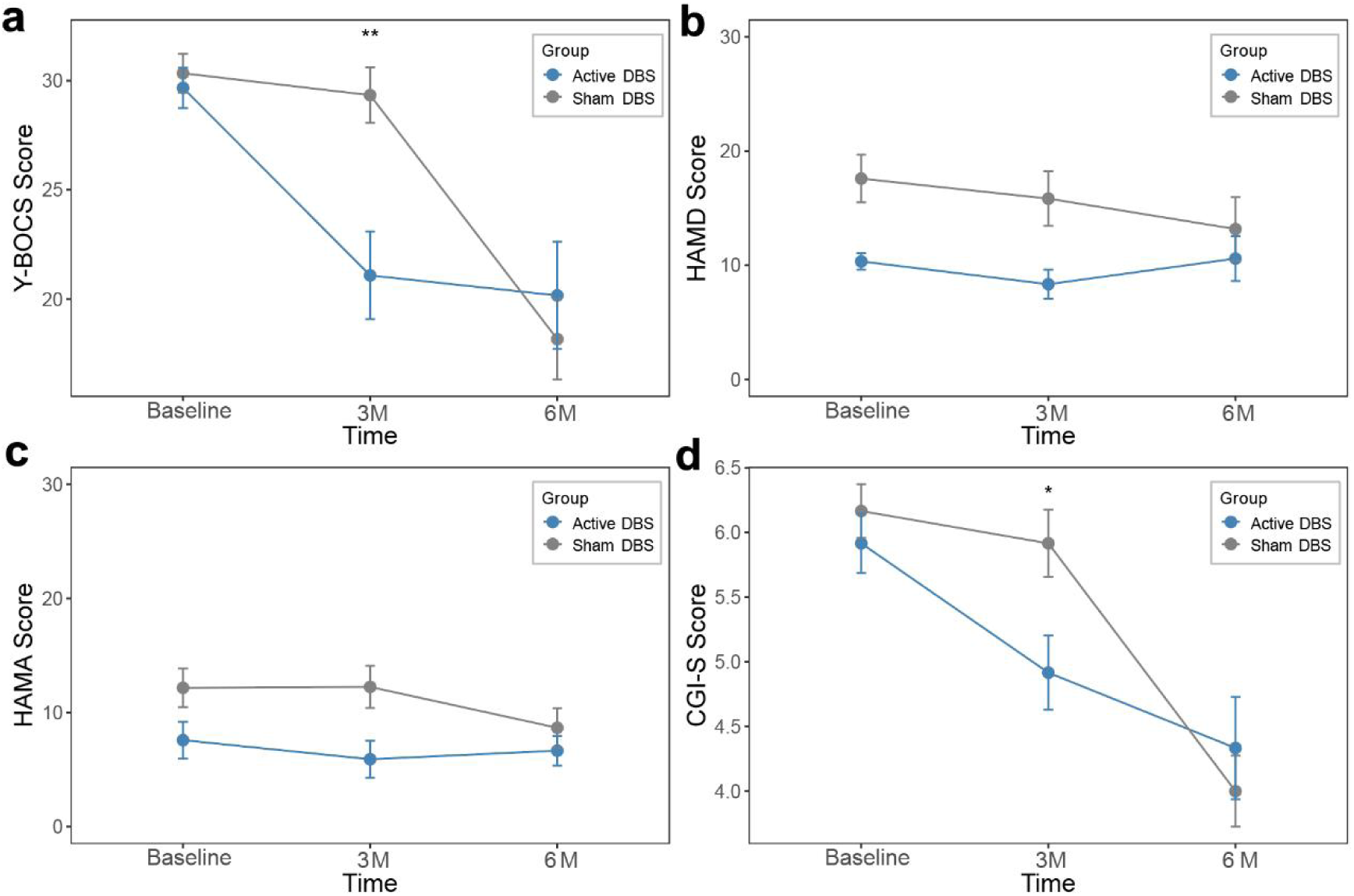
Longitudinal changes in clinical symptom scores in active versus sham DBS groups. **a**, Yale–Brown Obsessive–Compulsive Scale (Y-BOCS). **b**, Hamilton Depression Rating Scale (HAMD). **c**, Hamilton Anxiety Rating Scale (HAMA). **d**, Clinical Global Impression–Severity (CGI-S). Scores were assessed at baseline, 3 months (3M), and 6 months (6M) in the active DBS (blue) and sham DBS (gray) groups. Data are shown as mean ± SEM (\**p* < 0.05, \*\**p* < 0.001).

### Machine learning identifies a preoperative resting-state EEG signature for predicting DBS treatment outcome

Spectral power features from EEG have shown promise as treatment predictors for psychiatric disorders, with relative power offering improved sensitivity by accounting for inter-subject variability^32–34^. Therefore, in the present analysis, preoperative resting-state EEG recordings were analyzed to quantify log-transformed relative power across five frequency bands (delta (1-4 Hz), theta (4-7 Hz), alpha (7-12 Hz), beta (12-30 Hz), and gamma (30-50 Hz)) for 61 channels (Fig. S2) under two conditions (eyes-closed (EC) and eyes-open (EO)), yielding 610 candidate features per subject.

We aimed to predict symptom change, defined as the 6-month reduction rate in Y-BOCS scores (i.e., after participants in both arms received active stimulation), using a signature derived from resting-state EEG relative power features. Given the small sample size of subjects who have usable EEG data in the discovery cohort (*N* = 22; see Methods), we chose to identify a single feature as the predictor, as a complex multivariate classifier would risk poor generalization. To ensure an unbiased evaluation of the prediction performance, we implemented a nested leave-one-subject-out cross-validation (LOSOCV) procedure (Fig. 3a), which strictly separates the process of feature selection (across 610 features) from the final assessment of the model’s predictive performance to prevent data leakage.

**Fig. 3.**
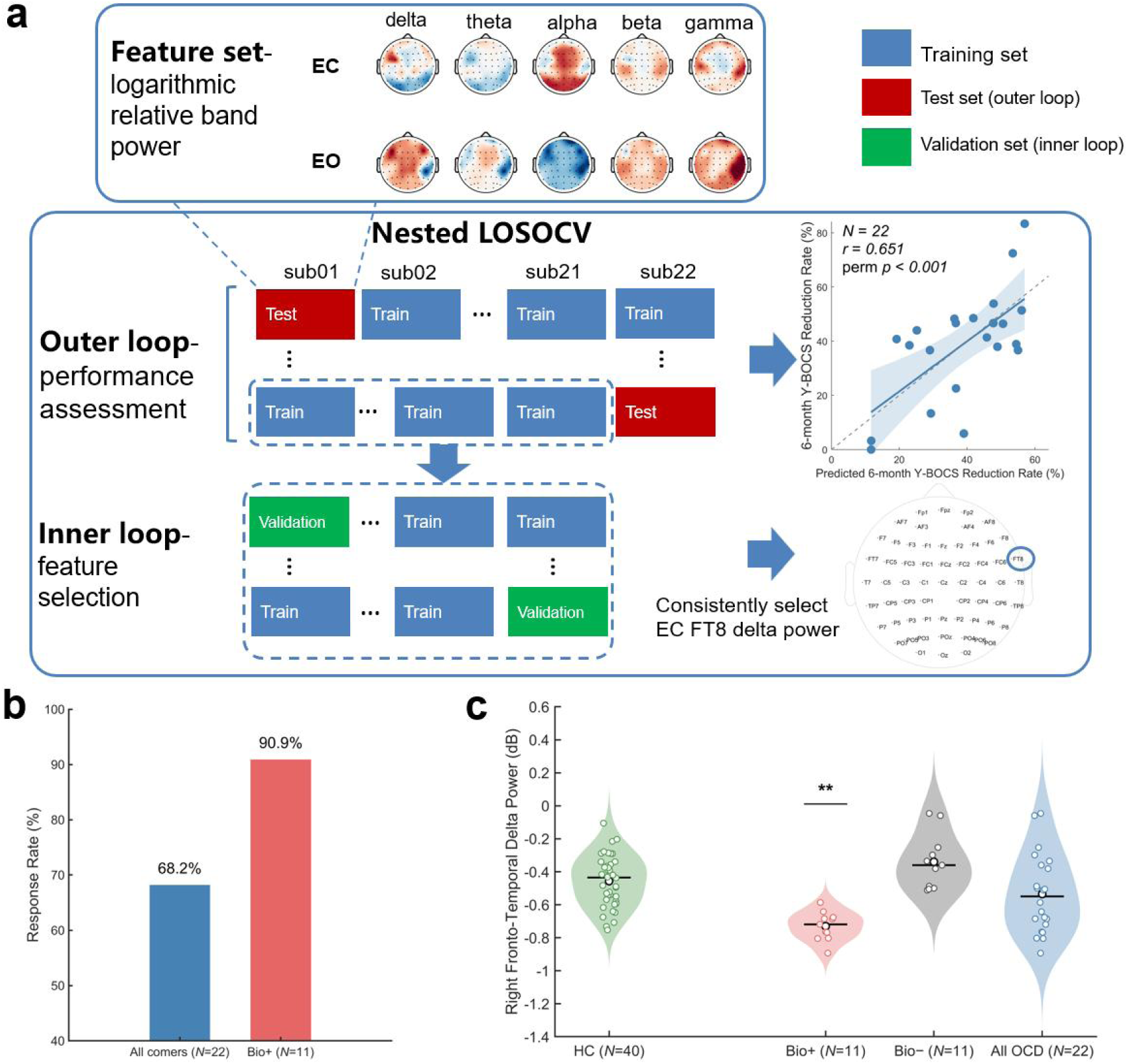
Identification of preoperative EEG treatment-predictive signature for DBS in the discovery cohort (*N* = 22). **a**, Prediction performance of the nested leave-one-subject-out cross-validation (LOSOCV) model for 6-month Y-BOCS reduction rate. Feature sets consisting of 610 features were derived from log-transformed relative power across five canonical frequency bands (delta, theta, alpha, beta, gamma) under EC and EO conditions, computed across all 61 EEG channels. The nested LOSOCV properly separates feature selection and performance assessment to prevent data leakage. An outer loop was used for model performance assessment, with one subject held out as the test set in each iteration and the remaining subjects split into training and validation sets via an inner loop for feature selection. Across iterations of the outer loop, EC FT8 delta power was consistently identified as the most predictive feature via the corresponding inner loops, showing a strong correlation with six-month Y-BOCS reduction rate (*r* = 0.651, 95% CI: 0.316 to 0.842; permutation *p* < 0.001). **b**, Response rates in all-comers (blue) versus biomarker-positive (Bio+) patients (red). **c**, Right fronto-temporal delta power in biomarker-defined subgroups and healthy controls. Bar plots with individual data points illustrate right fronto-temporal delta relative power (dB) in HCs (*N* = 40), biomarker-positive patients (Bio+, *N* = 11), biomarker-negative patients (Bio−, *N* = 11), and the combined OCD cohort (All OCD, *N* = 22). Error bars denote standard error of the mean (SEM). Welch’s *t*-tests were performed for pairwise comparisons between HC and each patient subgroup. Significance levels are indicated above the bars. ** indicates *p* < 0.001.

Across the 22 outer-loop iterations in nested LOSOCV, EC FT8 delta relative power was consistently selected as the optimal feature across all 22 iterations (Fig. S3). This complete consistency of feature selection argues against overfitting or spurious selection despite the modest sample size. In the outer loop, out-of-sample model predictions were significantly correlated with the observed 6-month Y-BOCS reduction rate (Pearson’s *r* = 0.651, 95% CI: 0.316 to 0.842, *R*^2^ = 0.421, *RMSE* = 0.1511, permutation *p <* 0.001; Fig. 3a), indicating a consistent association with treatment outcome.

Based on this finding, further analysis was performed to assess the direct association between EC FT8 delta relative power and symptom change on the full sample. Here again, this feature showed the strongest negative correlation with 6-month Y-BOCS reduction rate (Pearson’s *r* = −0.722, 95% CI: −0.877 to −0.432, permutation *p* < 0.001; Fig. S4), exceeding the strength observed at all other electrodes within the delta band under the EC condition (Fig. S5). This indicates that lower preoperative delta-band power in the right fronto-temporal region (hereafter referred to as *right fronto-temporal delta power*) predicts greater symptom improvement following treatment.

To enable patient stratification in clinical settings, an optimal cutoff for the treatment-predictive signature was determined in the discovery cohort using the Youden index^35^, which yielded a value of -0.586 (Fig. S4). This cutoff stratified individuals into a biomarker-positive (Bio+) group, defined as having preoperative right fronto-temporal delta power below this value, and a biomarker-negative (Bio-) group, defined as having preoperative right fronto-temporal delta power above this value. The Bio+ group exhibited a markedly higher response rate compared with the full cohort (90.9% vs. 68.2% ; Fig. 3b) and showed robust predictive performance, with a positive predictive value (PPV) of 90.9%, specificity 85.7%, sensitivity 66.7%, and normalized PPV (nPPV, defined as relative improvement over total PPV^36^) of 33.3%.

Compared to the full cohort, the Bio+ group showed a more pronounced difference in Y-BOCS and CGI-S scores between the active and sham DBS groups at the 3-month follow-up, with a substantially larger effect size (Y-BOCS: 95% CI: 10.31 to 15.80, Cohen’s d *=* 7.73, *p* < 0.001, CGI-S: 95% CI: 0.37 to 1.74, Cohen’s d *=* 2.51, *p* = 0.006). The Bio+ group showed no significant between-group differences in HAMA and HAMD score changes at the 3-month follow-up, consistent with the findings observed in the full cohort.

Furthermore, group comparisons of the EEG signature revealed that the Bio+ group exhibited significantly lower relative delta power than HCs (Welch’s *t* = 7.616, *p* <0.001; Fig. 3c), whose values were comparable to those observed in the Bio-group (Welch’s *t* = -2.086, *p =* 0.005). Concordant patterns were observed in both the FT8 delta spectral analysis (Fig. S6) and topographical maps of group-level log relative power analysis (Fig. S7). This divergence indicates that the Bio+ group displays a more pronounced deviation from normal EEG patterns than the Bio-group, suggesting a more severe underlying neurophysiological phenotype as reflected in its distinct EEG pattern.

### EEG signature is independent of baseline symptoms and demographics

To rule out the possibility that demographic factors or baseline severity mediated the predictive utility of the EEG signature; we examined whether it was associated with demographic or baseline clinical variables. This included age, sex, education years, BMI, course of illness, and baseline symptom severity scores (Y-BOCS total, obsession, compulsion, CGI-S, HAMD, and HAMA). Correlation analyses revealed no significant associations between the biomarker and any of these measures (all *p* > 0.336; table S1). Correlation coefficients ranged from –0.186 to 0.215, and none approached statistical significance.

### EEG signature specifically predicts clinical improvement under active DBS vis-à-vis sham DBS

A true treatment-predictive signature should be specific to the active treatment (i.e., it should moderate between outcomes with the active versus sham treatments) and not simply reflect a general tendency for improvement. To assess treatment specificity of the signature, we compared the correlation between right fronto-temporal delta power and 3-month Y-BOCS reduction rate following the double-blind phase in the active DBS group (*N* =10) and sham DBS group (*N* =12) of the discovery cohort. A significant negative correlation was observed in the active DBS group (Pearson’s *r* = −0.816, 95% CI: −0.955 to −0.383, permutation *p* = 0.007; Fig. 4a), whereas no comparable trend was detected in the sham DBS group (Pearson’s *r* = 0.168, 95% CI: −0.449 to 0.677, permutation *p* = 0.626; Fig. 4b).

**Fig. 4.**
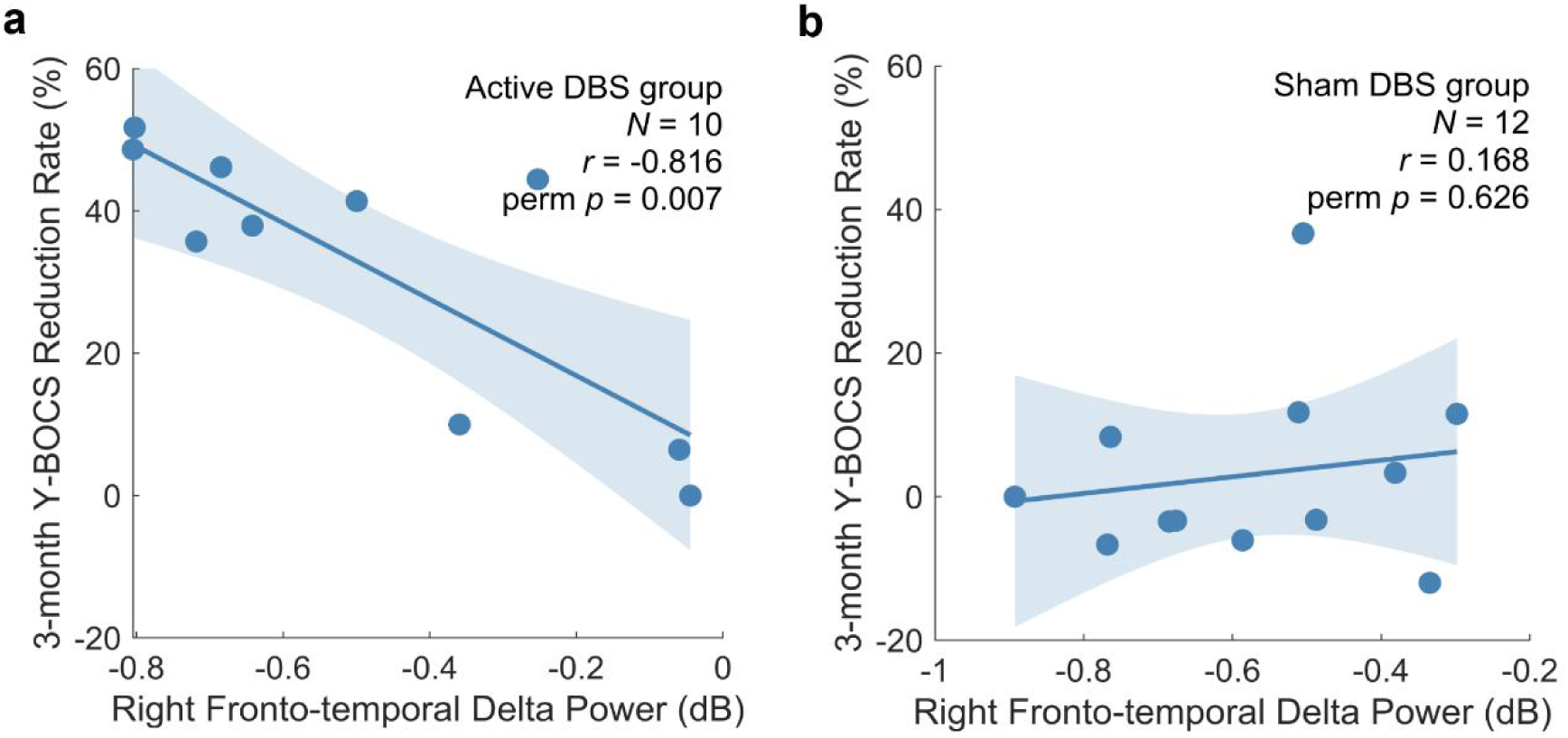
Specificity of the EEG signature to active DBS vis-à-vis sham DBS. Association between preoperative right fronto-temporal delta relative power and 6-month Y-BOCS reduction rate in the active DBS group (**a**; *N* = 10) and sham DBS group (**b**; *N* = 12), respectively. Solid blue line represents the regression fit, with shaded area denoting the 95% confidence interval.

### EEG signature prospectively predicts outcomes in an independent validation set

To assess its generalizability and clinical applicability, the predictive performance of the identified signature was prospectively evaluated in an independent validation set comprising 8 patients from cohorts 2 and 3 (ID T01–T08). These two cohorts were recruited after the treatment-predictive signature and its optimal cutoff value were finalized based on the results from the discovery cohort. Preoperative EC resting-state EEG was acquired, followed by standard DBS treatment and evaluation of Y-BOCS reduction rate at 3 months postoperatively.

The prediction model was based on right fronto-temporal delta power using a linear regression model (Fig. S4) and applied with a Youden index threshold to classify responders and non-responders, all derived from the discovery cohort. This model was locked and applied to the validation set without any post-hoc refinement. Model predictions were compared with clinician-assessed outcomes (Fig. 6a). Responder status was correctly classified in 7 of 8 patients (accuracy 87.5%; sensitivity 50.0% [1/2], specificity 100% [6/6], positive predictive value 100% [1/1], negative predictive value 85.7% [6/7]). The biomarker showed high specificity for identifying non-responders, whereas sensitivity was limited by the small number of responders, i.e., 2, in this cohort. Predicted and observed Y-BOCS reduction rates were significantly correlated across all subjects, when the validation predictions were pooled with the cross-validated discovery predictions (Pearson’s *r* = 0.620, permutation *p* < 0.001; Fig. 6b).

### EEG signature generalizes across resting-state conditions

To assess dependency of the EEG signature on resting-state conditions, right fronto-temporal delta power at electrode FT8 was also analyzed under the EO condition in the discovery cohort. The feature remained significantly negatively correlated with the 6-month Y- BOCS reduction rate (Pearson’s *r* = −0.498, 95% CI: −0.760 to −0.097, one-tailed permutation *p* = 0.008; Fig. 5a), consistent in direction with the EC condition, albeit with lower correlation strength. This consistency across resting-state conditions suggests that our signature reflects a stable neural state associated with DBS treatment outcome rather than a task-specific finding.

**Fig. 5.**
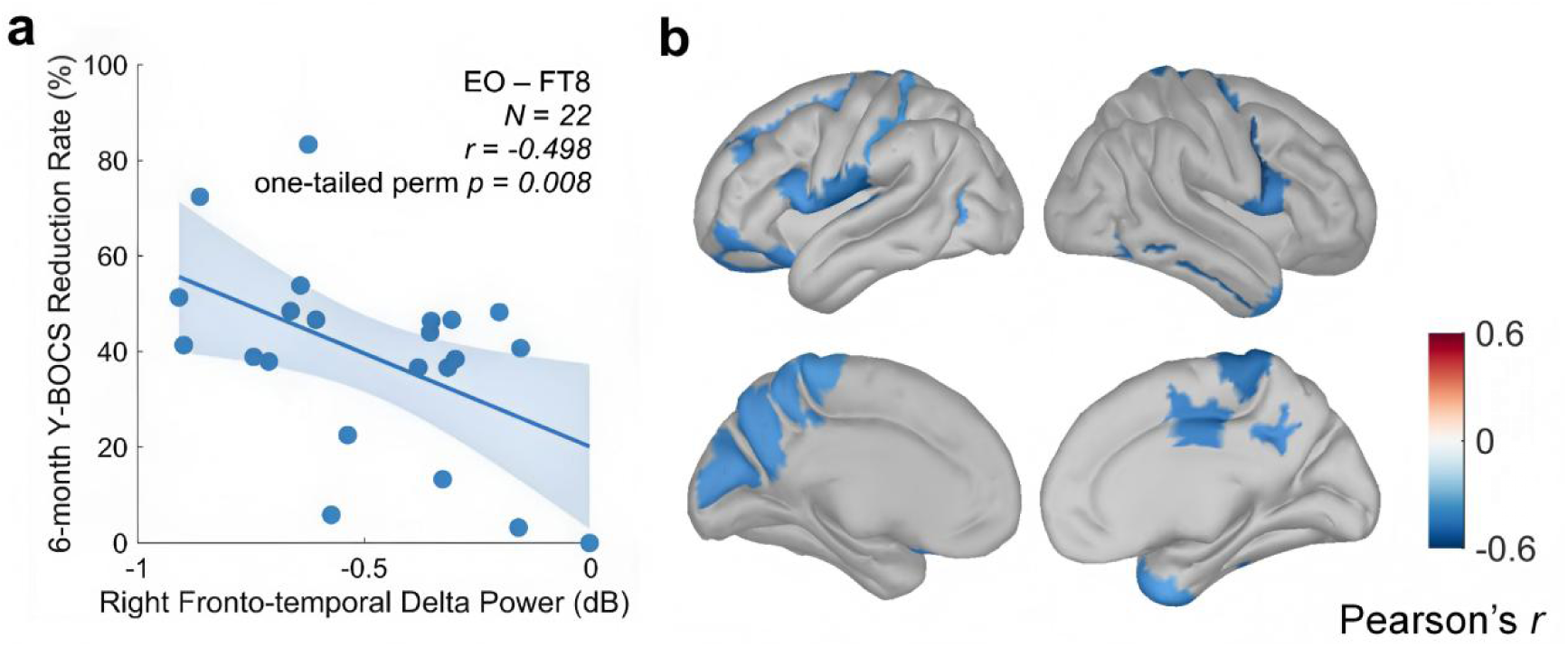
Correlation between preoperative fronto-temporal delta power and clinical outcome in an independent task paradigm (EO resting-state EEG) and modality (EC resting-state MEG). **a**, EO condition: right fronto-temporal (FT8) delta-band power versus 6-month Y-BOCS reduction rate. Solid blue line represents the regression fit, with shaded area denoting the 95% confidence interval. **b**, Cortical map of significant source-level correlations between delta-band power and 6-month Y-BOCS reduction rate in MEG. Significant ROIs identified by permutation testing (*p* < 0.05) are displayed on the cortical surface. All clusters showed negative associations (blue).

**Fig. 6.**
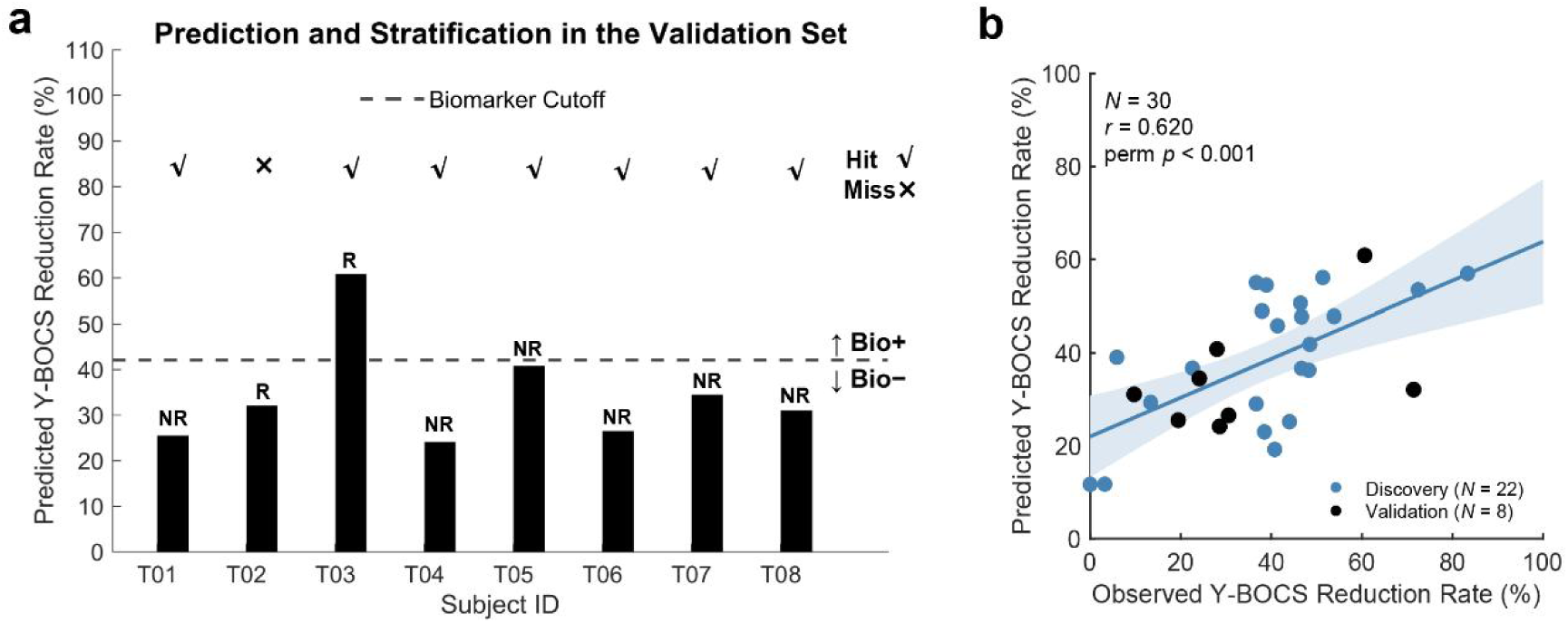
Model prediction and observed clinical outcomes in the external validation set. **a,** Predicted Y-BOCS reduction rate and stratification based on the right fronto-temporal delta power in 8 independent patients. The dashed line indicates the biomarker threshold (-0.586; determined using a Youden index threshold) used to distinguish Bio+ and Bio− individuals. Each patient’s true clinical outcome is labelled above the bars: R, responder; NR, non-responder. Check marks denote correct predictions, and crosses denote mismatches. **b**, Scatter plot of predicted versus observed Y-BOCS reduction rate across all subjects (*N*= 30), comprising the 22 discovery-cohort patients (blue points) and the 8 independent validation patients (dark points). Each dot represents one subject; the solid blue line indicates the linear fit and the shaded blue area the 95% confidence interval. Predicted and observed reduction rates were significantly correlated across all subjects (Pearson’s *r* = 0.620, 95% CI 0.335 to 0.801, permutation *p* < 0.001).

### MEG corroborates right frontotemporal predictive pattern

Since both EEG and MEG directly measure the brain’s underlying electrical activity, MEG provides an ideal complementary modality to validate our findings. To assess cross-modal robustness of the EEG signature, resting-state EC MEG data of the discovery cohort were source-localized onto individual cortices using participants’ T1-weighted MRI (*N* =19 patients with MEG data collected). Delta power was extracted and parcellated into 148 regions of interest (ROIs) with the Destrieux atlas^37^. Correlations were computed between each ROI’s delta power and 6-month Y-BOCS percent reduction rate, and statistical significance was evaluated with nonparametric permutation testing. All significant ROIs (*p* < 0.05) showed negative associations (Fig. 5b). The strongest effect occurred in the right inferior part of the precentral sulcus (S_precentral-inf-part R; Pearson’s *r = –*0.637, 95% CI: −0.846 to −0.257, one-tailed permutation *p =* 0.003; Fig. S8a), with an additional significant cluster in the opercular division of the right inferior frontal gyrus (G_front inf-Opercular R; Pearson’s *r = –*0.527, 95% CI: −0.792 to −0.096, one-tailed permutation *p =* 0.012; Fig. S8b). Anatomically, the latter falls within the ventrolateral prefrontal cortex (vlPFC), while the former lies immediately posterior to vlPFC near the inferior frontal junction (IFJ), a control hub at the intersection of the inferior frontal and precentral sulci implicated in cognitive control. The topography is also spatially consistent with the right fronto-temporal coverage of the FT8 EEG site reported in MRI-based cranio-cerebral correlation studies^38^, supporting convergence across modalities.

### EEG signature shows short- and long-term test-retest reliability

For a treatment-predictive signature to be clinically useful, it must be stable over time. To evaluate the stability of EC right fronto-temporal delta power as a treatment-predictive signature, its short-term and long-term test–retest reliability was assessed across resting-state EEG recordings from multiple cohorts using the intraclass correlation coefficient (ICC)^39^. Specifically, the short-term reliability was evaluated in two distinct groups. In the OCD discovery cohort (*N* = 21 patients with both preoperative and postoperative EEG data collected), the signature was consistent between pre- and postoperative recordings acquired before stimulation was initiated, separated by 1-3 months (ICC = 0.732; 95% CI: 0.506 to 0.864; Fig. S9a). This stability was confirmed in a healthy control dataset (*N* = 39) assessed two weeks apart (ICC = 0.686; 95% CI: 0.515 to 0.804; Fig. S9b). Long-term stability was demonstrated in a large, five-year healthy control dataset (*N* = 208). The signature’s reliability remained high in resting-state recordings acquired both before an intervening cognitive task (ICC = 0.667; 95% CI: 0.575 to 0.745) and after (ICC = 0.686; 95% CI: 0.602 to 0.755; Fig. S9c).

### EEG signature is associated with altered excitation–inhibition balance

To further probe the physiological basis of the right fronto-temporal EEG signature, we analyzed the aperiodic exponent at FT8 in the discovery cohort. The aperiodic exponent has emerged as a putative electrophysiological marker of cortical excitation–inhibition (E/I) balance, with lower values generally reflecting a shift toward increased excitation relative to inhibition. The right fronto-temporal aperiodic exponent was strongly correlated with baseline right fronto-temporal delta power (Pearson’s *r* = 0.816, 95% CI: 0.601 to 0.921, permutation *p* < 0.001; *N* = 22; Fig. 7a), indicating that the delta-power signature largely reflects variation in the aperiodic component of the EEG spectrum. When patients were stratified by biomarker status, Bio+ patients showed a significantly lower exponent than Bio− patients (Welch’s t test, *p* < 0.001; Fig. 7b)

**Fig. 7.**
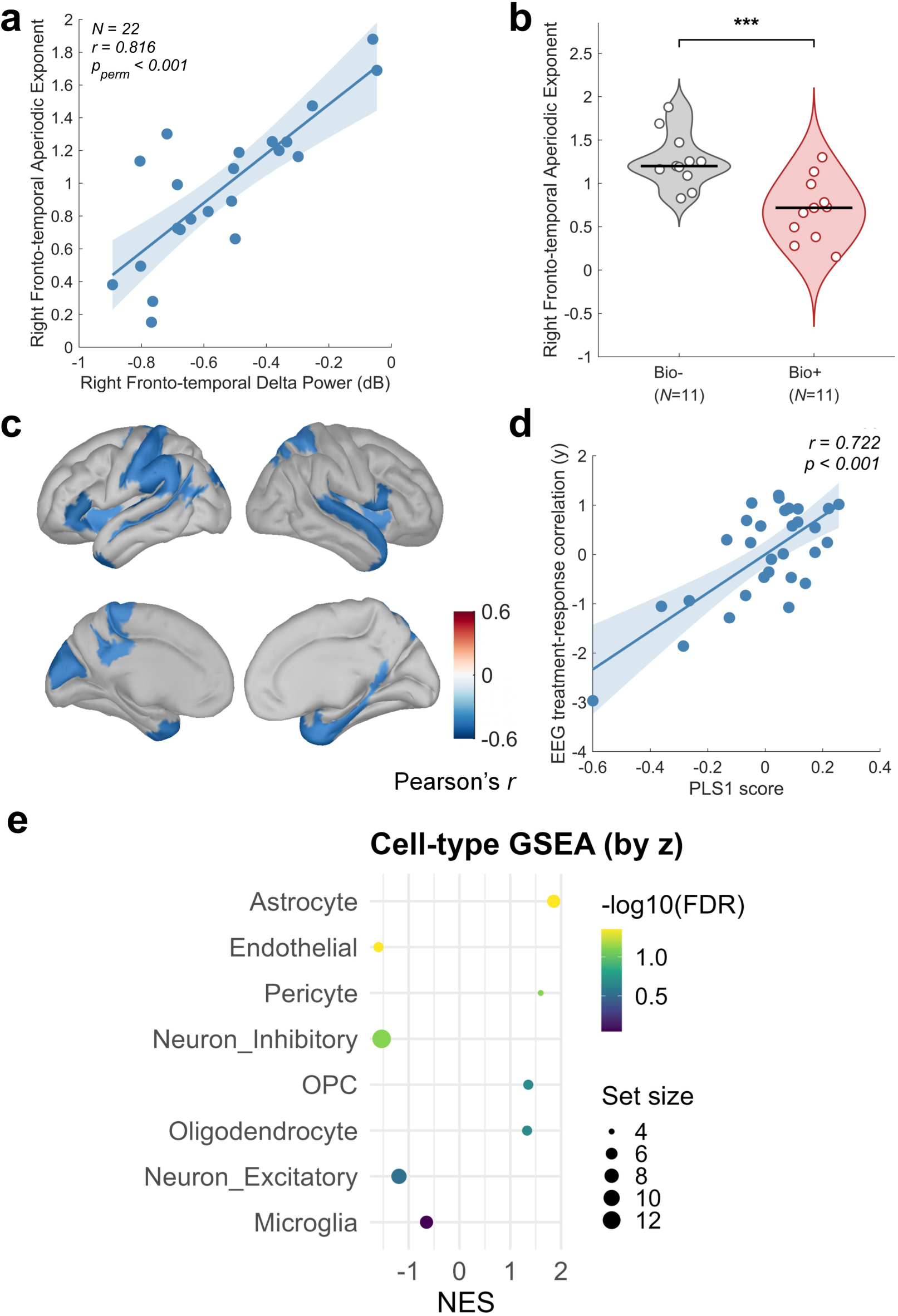
Mechanistic characterization of the EEG treatment-predictive signature through aperiodic analyses and transcriptomic. **a**, Correlation between baseline right fronto-temporal delta power and the aperiodic exponent at FT8 (each point is a patient, *N* = 22; line = linear fit, shaded band = 95% CI; Pearson’s *r* = 0.816, 95% CI: 0.601 to 0.921, permutation *p* < 0.001). **b**, Aperiodic exponent stratified by EEG-defined biomarker status; violins show the kernel-density distribution with individual patients (open circles) and the group median (horizontal line). Bio+ patients (pink, *N* = 11) had a lower exponent than Bio− patients (gray, *N* = 11; Welch’s t test, t(19.7) = −4.00, *p* < 0.001; \*\*\**p* < 0.001). **c**, Cortical map of correlations between delta-band power and 6-month Y-BOCS reduction rate in EEG. For each Destrieux cortical ROI, baseline delta-band relative power was correlated with 6-month Y-BOCS reduction rate after DBS treatment, and the surface map shows the 30 most strongly negatively correlated ROIs; **d**, PLS analysis relating source-space EEG treatment-predictive pattern to cortical gene expression, showing the first latent axis (PLS1): each point is a cortical region, the y-axis is the EEG treatment-predictive strength, the x-axis is the corresponding PLS1 score, and the fitted line illustrates their positive association (Pearson’s *r* = 0.722, 95% CI: 0.635 to 0.791, *p*<0.001); **e**, Cell-type GSEA (broad classes: Astrocyte, Neuron_Inhibitory, Microglia, Endothelial, Neuron_Excitatory, OPC, Pericyte, Oligodendrocyte). Points encode NES on the x-axis, −log10(FDR) by color, and gene-set size by dot size. Neuron_Inhibitory is significant (NES = -1.530, *p* = 0.008, FDR = 0.033, set size = 13).

### DBS-treatment-predictive pattern in EEG is linked to inhibitory-neuron marker expression

To explore the microscale biological basis of the predictive EEG pattern associated with the DBS treatment, we investigated its relationship with cortical gene expression. This analysis sought to determine whether the cortical regions where baseline rsEEG delta-band power is predictive of the DBS treatment align with specific cellular transcriptional programs.

First, we created a source-space EEG-treatment association map by correlating the baseline delta-band power in each source-space ROI with the 6-month Y-BOCS reduction rate. The resulting map (Fig. 7c) identifies the cortical areas where baseline delta power was associated with clinical improvement, where the 30 most strongly negatively correlated ROIs are displayed. The spatial pattern showed exclusively negative correlations, indicating that lower baseline delta power predicted greater 6-month Y-BOCS reduction rate after DBS. This topography was largely consistent with the EEG sensor-level findings, with pronounced effects in the bilateral temporal and orbitofrontal cortices, as well as the left parietal regions. We then used PLS regression to find the dominant axis of cortical gene expression (PLS1) from the Allen Human Brain Atlas (AHBA) that spatially mirrored this association map. We found a significant positive correlation between a region’s gene expression profile (PLS1, permutation *p* = 0.046) and its score on the EEG-treatment association map (Pearson’s *r* = 0.722, 95% CI: 0.635 to 0.791, *p* < 0.001; Fig. 7d). In other words, cortical areas whose transcriptional profile loads more strongly onto PLS1 are the same areas whose baseline delta-band activity is more negatively predictive of subsequent clinical improvement.

To explore the cellular correlates of this association, we used the PLS1 gene ranking to perform a pre-ranked Gene Set Enrichment Analysis (GSEA). This analysis revealed that the gene profile was significantly and negatively enriched for inhibitory-neuron markers (NES = -1.53, FDR = 0.033; set size = 13), Fig. 7e). Given that the association map is positively correlated with the PLS1 gene score, the negative enrichment for inhibitory-neuron markers at the high-PLS1 end indicates that expression of inhibitory neurons is highest in regions that are more negatively predictive, suggesting an inherent molecular deficit in inhibitory signaling of these regions.

### DBS-induced changes in EEG signature track clinical improvement

While a treatment-predictive signature need not be modifiable by treatment, understanding whether DBS modulates the identified signature can reveal its potential as a physiological readout of target engagement. To examine whether DBS modulates the right fronto-temporal delta power signature and whether such changes track clinical response, we related changes in the right fronto-temporal delta power signature over the first 3 months of stimulation (baseline-to-3-month for the active group and 3-to-6-month for the sham group) to the Y-BOCS reduction rate after 3 months of DBS in the discovery cohort. This alignment ensured that signature changes and symptom improvements were temporally matched. Specifically, the signature percent change was computed as the percentage change in the 3-month delta power signature relative to its earlier reference value. To limit the influence of extreme values, the signature percent change was winsorized to −100% to 100%. Across 19 participants with EEG data collected at the follow-up timepoints, greater increases in relative delta power were associated with stronger clinical improvement (Pearson’s *r* = 0.475, 95% CI: 0.027 to 0.764, RMSE = 0.171, one-tailed permutation *p* = 0.019; Fig. 8a). When stratified by EEG-defined biomarker status (Fig. 8b), Bio+ patients (*N* = 11) showed a substantially larger increase in relative delta power over the treatment interval than Bio− patients (*N* = 8; Welch’s t-test, *p* = 0.030).

**Fig. 8.**
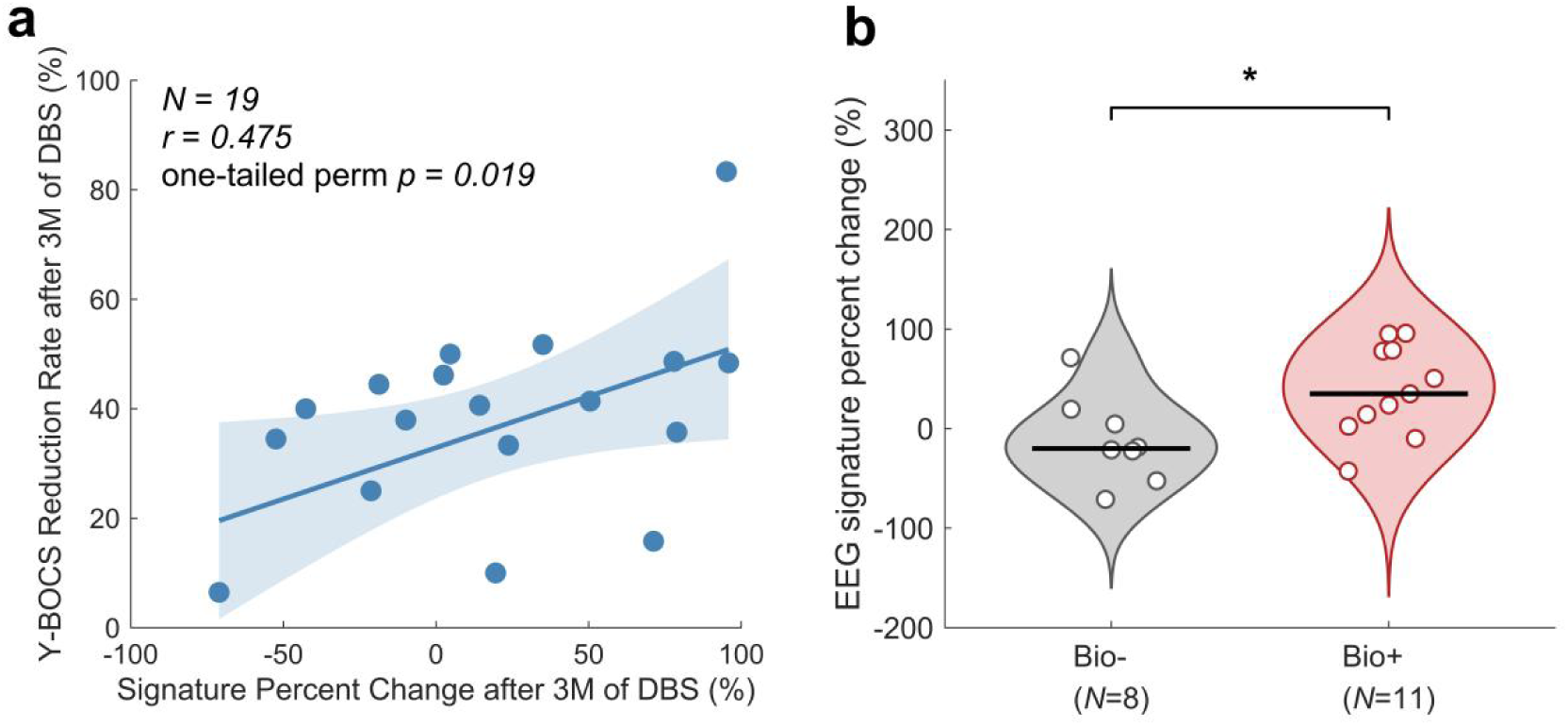
Longitudinal changes in right fronto-temporal delta power track clinical improvement after DBS. a, Scatter plot of right fronto-temporal delta power percent change vs. Y-BOCS reduction rate after 3 months of DBS in 19 participants with follow-up (Pearson’s *r* = 0.475, 95% CI: 0.027 to 0.764, one-tailed permutation *p* = 0.019). b, Percentage change in relative delta power stratified by EEG-defined biomarker status. Violins show the kernel-density distribution with individual participants (open circles) and the group median (horizontal line); Bio+ patients (pink, *N* = 11) showed a greater increase than Bio− patients (gray, *N* = 8) (Welch’s t-test, 95% CI: 5.449 to 93.751%, *p* = 0.030; \**p* < 0.05).

## Discussion

This study demonstrated that dual-target DBS for ALIC and NAcc significantly alleviated symptoms in patients with treatment-refractory OCD. At the 3-month follow-up, the active DBS group showed significantly greater improvement in Y-BOCS reduction rate and CGI-S scores compared to the sham stimulation group (Fig. 2), consistent with the clinical specificity and efficacy of this treatment.

Building on this dual-target therapeutic paradigm, we identified and prospectively validated a preoperative, non-invasive EEG signature that predicted the clinical response to DBS in OCD. Through nested LOSOCV that prevented data leakage between feature selection and assessment of prediction performance, we found that lower relative delta power in the right fronto-temporal region, measured from EC resting-state EEG before surgery, was strongly predictive of greater symptom improvement following active DBS (Fig. S4). This finding has direct clinical relevance, as such an indicator could help optimize patient selection, improve overall response rates (Fig. 3b), and reduce the risk of unnecessary invasive procedures for likely non-responders.

Decreased resting-state relative delta power, particularly over frontal regions, was reported by prior studies in non-surgical OCD cohorts and interpreted as a marker of cortical dysfunction^42^. Our results refine this understanding by indicating that this decrease is driven specifically by the Bio+ subgroup; neither Bio− patients nor the OCD cohort as a whole differed significantly from HCs (Fig. 3c). The hyperactivity of slow-wave oscillations is thought to be a downstream consequence of a more fundamental excitation/inhibition (E/I) imbalance within the CSTC circuits^22^. Our results suggest that patients with less pronounced baseline slow-wave cortical dysfunction—that is, those with lower preoperative delta power—may possess a greater capacity for their neural circuits to be favorably modulated by DBS.

The sham-controlled design of the RCT allowed us to establish the treatment specificity of this EEG signature. The predictive relationship between lower preoperative right fronto-temporal delta power and greater symptom reduction was present only in the active DBS group and was absent in the sham DBS group (Fig. 4). This finding suggests that the signature is not merely a general prognostic indicator of a more favorable disease course but is specifically linked to the patient’s capacity to respond to the neurophysiological effects of active DBS. This specificity supports the signature’s validity as a true predictor of treatment response.

For any treatment-predictive signature to be clinically useful, it must be both reliable and robust. We established the stability of right fronto-temporal delta power through multiple analyses. The signature showed good short-term test-retest reliability within our patient cohort (pre- and postoperative recordings acquired before stimulation was initiated) and excellent long-term reliability in a large, independent dataset spanning five years (Fig. S9). Furthermore, its predictive power was consistent across both EC and EO resting-state conditions (Fig. 5a), suggesting it reflects a stable, trait-like neural feature rather than a transient state. The robustness of the signature was further underscored by its cross-modal consistency between EEG and MEG (Fig. 5b). Given the significant differences in their physical measurement mechanisms—MEG being more sensitive to tangential neural currents, while EEG is more influenced by the conductive properties of head tissues—the ability to reproduce this finding across both modalities provides strong evidence for its biological plausibility, corroborating that the biomarker reflects intrinsic neural activity in the fronto-temporal cortex and is not an artifact of a single measurement technique.

The most stringent test of a treatment-predictive signature is its ability to generalize to new, unseen data. Our signature successfully predicted clinical outcomes with high accuracy in a completely independent, prospectively recruited validation cohort. Specifically, the signature successfully predicted the response status of 7 out of 8 patients, and the predicted Y-BOCS reduction rates were highly consistent with the actual reduction rates (Fig. 6). This successful out-of-sample validation is a critical step toward establishing the signature’s potential for clinical translation and supports its potential generalizability beyond the initial discovery sample.

Beyond its predictive utility, we also found preliminary evidence that our EEG signature may track treatment response (Fig. 8), this finding should be regarded as proof of concept rather than definitive evidence. The ability to track therapeutic efficacy is of great clinical significance as it offers a potential readout for target engagement^28^. While traditional clinical scales such as Y-BOCS and CGI-S remain gold standards for efficacy assessment, they require manual interviews and subjective scoring, are time-consuming, depend on patient cooperation, and may lack sensitivity for short-term monitoring. In contrast, an objective, EEG-based neurophysiological indicator offers the advantages of being rapid, objective, and repeatable, providing quantitative information on neural functional status to complement clinical follow-up. Our finding that changes in right fronto-temporal delta power over the course of active DBS group were significantly correlated with the degree of clinical improvement brings us closer to identifying such an indicator. Specifically, responders showed a significant increase in delta power during the treatment period, moving toward the levels seen in preoperative non-responders. Our results align with the work of Smith et al.^25^, which found that increases in prefrontal delta power during active VC/VS DBS correlated with better outcomes. Furthermore, a recent study^41^ investigating biomarkers for cognitive behavioral therapy (CBT) in depression reported that a better response to CBT was predicted by lower baseline delta power, and that responders showed a significant increase in delta power after just two weeks of therapy. The existence of this parallel pattern—that resting-state delta activity predicts and tracks response to two markedly different circuit-based interventions (DBS and CBT) in two different disorders (OCD and depression)—suggests that this EEG signature may be a transdiagnostic marker of a brain state that is highly amenable to therapeutic regulation. Ultimately, the signature’s ability to track therapeutic progress suggests it may reflect the state of a causally relevant brain circuit^14^, which can potentially be leveraged to guide the optimization of stimulation parameters and aid the development of future closed-loop DBS systems for OCD.

Our identification of right fronto-temporal delta power as the treatment-predictive signature provides insight into the inhibitory control network that prior studies have shown to mediate DBS therapeutic effects. The right fronto-temporal region is a key hub in circuits responsible for emotion regulation, impulse control, and reward processing—functional domains known to be central to the pathophysiology of OCD^43,44^. In particular, Li et al. provided compelling connectomic evidence that multiple effective DBS targets for OCD—including the ALIC, NAcc, and VC/VS—converge on a common ultra-fast fiber pathway to the vlPFC^15^. Converging evidence from functional neuroimaging, lesion, and stimulation studies indicates that the vlPFC—particularly its right inferior frontal gyrus (rIFG) subregion—is a crucial cortical region for inhibitory control, robustly activated during stop-signal and related response inhibition tasks^45^. Activation of this pathway is therefore well-positioned to engage top-down inhibitory control over downstream CSTC loops, which are critically implicated in compulsive symptom generation^46^. This right-lateralized inhibitory network, encompassing the rIFG and adjacent vlPFC, has been consistently associated with superior clinical outcomes when effectively engaged by stimulation fields^15^. In our study, right fronto-temporal delta power—captured at FT8—might be a robust, non-invasive electrophysiological marker for the brain’s top-down control signal originating from this inhibitory network. Lower preoperative delta power may reflect severe deficit in this top-down control, indicating a hyper-excitable and less regulated CSTC circuit. This greater neurophysiological dysfunction, however, also signifies greater capacity for DBS-induced restoration of inhibitory control, explaining why patients with lower baseline delta power exhibited greater symptom improvement after combined ALIC and NAcc DBS. Furthermore, the observed post-treatment increase in delta power among responders is consistent with the successful re-engagement of this top-down control system via the ALIC–vlPFC pathway, aligning with Li et al.’s connectomic model^15^.

Mechanistically, the aperiodic exponent analysis provided complementary support for the link between the EEG signature and baseline excitation–inhibition balance. The right fronto-temporal aperiodic exponent at FT8 was strongly correlated with the EEG signature and was significantly lower in Bio+ than in Bio− patients (Fig. 7), indicating that the signature largely tracks the aperiodic slope. Because a flatter (lower-exponent) spectral slope has been linked to relatively greater excitation over inhibition, this suggests that the FT8 delta-power signature may partly reflect an altered E/I-related cortical state relevant to DBS response. Consistent with this interpretation, our transcriptomic association analysis that investigated the relationship between the treatment-predictive strength of EEG and its underlying transcriptional architecture established that the cortical regions with the strongest predictive power were those with the highest expression of these inhibitory-neuron markers (Fig. 7), lending support to the involvement of these cortical regions in inhibitory control.

Importantly, resting-state EEG as a recording modality offers inherent advantages for clinical translation: it is inexpensive, non-invasive, and readily available in many clinical settings. Unlike task-based EEG paradigms that require specific cognitive engagement and is more demanding in terms of patient cooperation, resting-state recording is straightforward to implement in routine practice, especially with portable systems. Building on the identified EEG signature, standardized protocols can be developed to automatically compute an EEG-derived signature value based on a brief, 5-minute recording, ensuring consistency and reproducibility across clinical sites. Such simplicity and cost-efficiency make resting-state EEG particularly suitable for large-scale adoption, enabling its integration into everyday psychiatric workflows and supporting biomarker-guided decision-making in patient selection for DBS.

Several limitations of this study should be noted. First, while our discovery set represents among the largest cohorts for an RCT of OCD DBS to date, the sample size is still limited for employing more sophisticated data-driven methods, such as machine learning approaches that aggregate information across eye conditions, electrodes, and frequency bands for biomarker identification. Similarly, our independent validation set, while providing crucial prospective evidence, was small. Additionally, the treatment-predictive signature was developed and validated using data collected from a single study site, which may limit its generalizability across different clinical settings, patient populations, or acquisition hardware. Therefore, further validation in larger, multi-site prospective cohorts is necessary to confirm its clinical utility. Moreover, while our findings link fronto-temporal delta power to the outcomes of NAcc and ALIC stimulation and provide convergent insights at the transcriptomic and behavioral levels, the precise neurophysiological mechanisms underlying this relationship remain to be fully elucidated. Future work combining computational modeling with neuroimaging or intracranial recordings could explore how baseline cortical states influence the network-level effects of deep brain stimulation^47^. Finally, it is currently unknown whether this signature could also predict outcomes for other DBS targets in OCD, such as the STN.

## Materials and Methods

### Study design and participants

Identification of the treatment-predictive EEG signature was conducted in Cohort 1, using data from a single center conducted from December 2021 to January 2024 at Shanghai Mental Health Center within a randomized, sham-controlled, double-blind, multicenter study (The Efficacy and Safety of ALIC/NAcc-DBS for Treatment-refractory OCD, NCT04967560). The protocol of this trial has been published and approved by the Human Research Ethics Committees of the Shanghai Mental Health Center (2021-15R4). This study consisted of three phases: (a) the screening and implantation phase, during which all participants underwent ALIC-NAc DBS surgery; (b) the three-month double-blind phase, in which patients were randomly assigned to receive either active or sham stimulation; and (c) the three-month open-label phase, during which all patients received active stimulation.

Participants aged 18–65 years with a primary diagnosis of OCD based on the Diagnostic and Statistical Manual of Mental Disorders, fifth edition were recruited. Other inclusion criteria included a minimum score of 25 on the Yale–Brown Obsessive-Compulsive Scale (Y–BOCS) accompanied by substantial functional impairment and meeting the criteria for treatment-refractory OCD. Treatment refractory was defined as a lack of response to: (a) at least three adequate pharmacological trials with first- and second-line medications (at least one trial using clomipramine), at or exceeding the maximum recommended dose; (b) augmentation with at least two antipsychotic drugs; and (c) sufficient trials of cognitive-behavioral therapy (at least 20 sessions of therapist-guided exposure and prevention therapy). Patients were excluded from the study if they met any of the following criteria: (a) have comorbid schizophrenia spectrum and other psychotic disorders, bipolar disorder, major depression disorder, etc.; (b) had severe neurological or physical illness; (c) had contraindications to neurosurgery; (e) had substance abuse or dependence; (f) were pregnant females; or (g) had severe suicide risk and tendencies.

Data for result validation were derived from Cohorts 2 and 3. Cohort 2 was an exploratory open-label study (Efficacy and Safety of Combo-stim Deep Brain Stimulation for Treatment-refractory Mental Disorders, NCT06112067) in which participants underwent ALIC-NAc DBS implantation followed by a 6-month follow-up period. Cohort 3 was a randomized, open-label, parallel-controlled study (NCT07031544) in which participants were randomly assigned to one of three groups based on DBS activation timing—one, two-, or 3-month post-surgery—and were followed for six months after stimulation onset. Both of two studies have been approved by the Human Research Ethics Committees of the Shanghai Mental Health Center (cohort 2:2022-91R3, cohort 3:2024-69R1). The inclusion and exclusion criteria were largely consistent with those of Cohort 1, except for the definition of treatment-refractory OCD (criterion a). In Cohorts 2 and 3, patients were required to have undergone adequate trials of at least three different selective serotonin reuptake inhibitors, without the mandatory inclusion of clomipramine.

All participants provided informed consent after receiving an explanation of the study procedure. All procedures contributing to this study comply with the ethical standards of the relevant national and institutional committees on human experimentation and with the Helsinki Declaration.

### DBS procedure

The placement of DBS electrodes was determined by a preoperative MRI scan with the Leksell surgical planning system (Surgiplan™, Elekta, Stockholm, Sweden). The insertion of custom tetrapolar electrodes (model 1242, SceneRay, Suzhou, China) along the trajectory of the ALIC extended into the NAc (Fig. S11). The lead contacts were 3.0 mm long and the contact spacing were 2.0 mm, 4.0 mm, and 4.0 mm, respectively, from ventral to dorsal. The electrode leads were externalized to confirm the electrode locations and perform a temporary stimulation. The NAc targets was set at a lateral distance of 7–12 mm from the midline, an anterior distance of 5–7 mm from the anterior border of the anterior commissure, and an inferior distance of 4–6 mm from the intercommissural line for reference. The two ventral contacts were preset with the NAc, while the two dorsal contacts were preset into the ALIC. A connecting wire (model SR1341, SceneRay, Suzhou China) from the scalp was connected to a subcutaneous implantable pulse generator (IPG; model SR1181, SceneRay, Suzhou, China). The IPG equipped with a non-rechargeable battery, was subcutaneously implanted in the right subclavicular area. Postoperative cranial computerized tomography (CT) scans were performed to ensure accurate electrode placement. After 2-4 weeks of recovery from implantation surgery, all participants underwent a programming procedure to determine individualized programming parameters.

### Clinical assessments

We make assessment of the obsessive-compulsive symptom severity by Yale-Brown Obsessive-Compulsive Scale (Y-BOCS), the anxiety symptom severity by the Hamilton Anxiety Scale (HAMA), the depressive symptom by the Hamilton Depression Scale (HAMD) and the overall clinical symptom severity by Clinical Global Impressions (CGI) scale including CGI-Severity (CGI-S) and CGI-Improvement (CGI-I) at baseline, 3-month follow-up and 6-month follow-up. The treatment response rate was defined as the number of patients who demonstrated a response to treatment, divided by the total number of participants in that group. Treatment response was defined as a more than 35% reduction rate in Y–BOCS score, and a CGI-I score of 2 or less.

### Data acquisition

Resting-state EEG was recorded for 5 minutes EO and 5 minutes EC state at baseline. Participants were seated in a separate room and remained awake during EEG recording. Data were collected using a 64-channel EEG system (Eemagine, Berlin, Germany), following the International 10–10 system, with CPz as reference and AFz as ground (the ground electrode was not included in the 64 recording channels). The sampling rate was 500 Hz, and electrode impedance (Ag/AgCl ring electrodes) was maintained below 10 kΩ.

MEG data were acquired using the Elekta Neuromag® TRIUX system (Elekta, Stockholm, Sweden), a helmet-shaped whole-head system comprising 102 sensor locations, each equipped with three sensors (one magnetometer and two gradiometers), totaling 306 channels. The sampling rate was 1000 Hz. During the recording, participants were instructed to keep their head and body stable while staying awake with their eyes naturally closed.

Structural MRI data were acquired using a Siemens® 3.0T Verio scanner. Participants were instructed to remain as still as possible, maintaining head and body stability while staying awake with eyes naturally closed, with active cooperation encouraged. Whole-brain structural images were obtained using an MPRAGE sequence with the following parameters: repetition time (TR) = 2300 ms, time echo (TE) = 3.5 ms, flip angle (FA) = 9°, field of view (FoV) = 256 mm, voxel size = 1.0 mm × 1.0 mm × 1.0 mm, matrix size = 256 × 176, slice gap = 0, and bandwidth = 180 Hz/px.

### Datasets

This study included multimodal data (EEG, MEG, MRI-T1, and fMRI) from three OCD patient cohorts for the discovery and validation of the treatment-predictive EEG signature. The test-retest reliability of the signature was then assessed using EEG data from two healthy control datasets used for evaluating the short-term and long-term reliability, respectively.

### Discovery set (OCD cohort 1)

In the biomarker discovery stage, 24 patients with OCD were initially recruited. Of these, 23 underwent preoperative resting-state EEG recordings. One patient was excluded because spectral quality was globally abnormal: all channels exhibited an aperiodic exponent > 3, and the mean exponent reached 5.263, far exceeding the physiologically plausible range (∼0.5–3.0 in EEG/MEG recordings^48^), leaving 22 valid baseline EC resting-state EEG datasets for biomarker discovery analyses.

For validation, two complementary datasets were used. First, preoperative EO resting-state EEG from the same 22 patients was analyzed to provide cross-condition validation. Second, 19 of these patients underwent preoperative resting-state MEG accompanied by high-quality structural T1-weighted MRI, which enabled MEG source-space reconstruction and cross-modal validation.

A postoperative pre-stimulation baseline EEG dataset was available for 21 patients. Together with their corresponding preoperative EEG, these 21 paired datasets were included in test–retest reliability analyses.

Longitudinal follow-up datasets were also obtained from the discovery cohort. At 3 months, EEG data were available for 22 patients (active DBS, *N* = 10; sham DBS, *N* = 12). At 6 months, EEG data were available for 19 patients (active DBS, *N* = 9; sham DBS, *N* = 10). These follow-up datasets were included in longitudinal analyses. Missing follow-up data were due to clinical scheduling differences and acquisition failures.

### Validation set (OCD cohorts 2 and 3)

The validation set included 8 patients from cohorts 2 (T01–T03) and 3 (T04–T08). All underwent the same preoperative EO and EC resting-state EEG protocol as cohort 1, as well as resting-state MEG recordings and T1-weighted MRI scans for MEG source modeling. All validation patients received three months of active stimulation.

### Healthy control dataset 1

Healthy control dataset 1 was obtained from a randomized controlled trial that included a healthy control group (Efficacy and Influencing Factors of Deep Transcranial Magnetic Stimulation in the Treatment of OCD, NCT06692114). EEG data were collected from 40 healthy control participants under identical laboratory conditions (same site, equipment, and personnel) and repeated two weeks later. One participant lacked Session 2 data, resulting in 39 subjects available for the test–retest analysis. This dataset was used to evaluate short-term test–retest reliability of the EEG spectral measures applied in the OCD cohorts. In addition, Session 1 data were used as the benchmark healthy control group for case–control comparisons with OCD patients. The study was approved by the Human Research Ethics Committees of the Shanghai Mental Health Center (2024-68), and all participants provided informed consent after receiving an explanation of the study procedures.

### Healthy control dataset 2

Long-term test–retest reliability was evaluated using a publicly available EEG dataset comprising 64-channel resting-state recordings (Openneuro accession number: ds005385)^49^. The dataset covers a wide adult age range with relatively even distribution and includes longitudinal data, enabling investigation of cross-sectional resting-state EEG patterns across the adult lifespan, assessment of cognitive activity and fatigue effects, and tracking of intra-individual changes over an interval of approximately five years. These data can also serve as a normative reference for clinical comparisons.

Resting-state EEG was recorded for 3 minutes each in EC and EO conditions. EEG was acquired both before and after an approximately 2-hour cognitive test battery designed to engage five domains of cognitive function: (i) visual attention, (ii) vigilance, (iii) conflict processing, (iv) updating and learning, and (v) speech perception (https://vital-study.ifado.de/measures/cognitive-tests-with-eeg-recording/tasks-on-day-1).

From the initial 608 participants (session 1), 208 completed a follow-up assessment approximately five years later (session 2). EEG was collected using a 64-channel elastic cap (10–20 system, FCz online reference), a BrainVision BrainAmp DC amplifier, and BrainVision Recorder software (BrainProducts GmbH). Signals were sampled at 1,000 Hz with a 250 Hz online low-pass filter, no online high-pass filter, and electrode impedances kept below 10 kΩ. The dataset is publicly available via Openneuro (accession number: ds005385, https://doi.org/10.18112/openneuro.ds005385.v1.0.2).

### Statistical analysis of clinical data

Clinical data analysis was conducted using SPSS (version 27.0, IBM Corp, NY, USA) and R Statistical Software (v4.4.3; R Core Team 2025). Statistical analyses of clinical data were conducted on the intention-to-treat (ITT) population, which included all participants who entered randomization. Missing values were imputed using the last observation carried forward method. In the ITT population, baseline demographic and clinical characteristics were summarized using percentages, means and SD, or medians and interquartile range (IQR; defined as the difference between the 75th and 25th percentiles), depending on the data distribution. A Fisher’s exact test was used to compare the response rates between the active DBS and sham DBS groups at the 3-month follow-up. A mixed linear model was used to compare the change in Y–BOCS, CGI, HAMD, and HAMA scores at the third and sixth months relative to baseline between the active DBS and sham DBS groups, with treatment, time, and the interaction between treatment and time as fixed effects, subjects as random effects, and baseline measurements as covariate variables. The estimated differences and 95% CI were calculated from the mixed model. Cohen’s d effect size was calculated by dividing the estimated fixed effect coefficient by the SD of the residuals.

### EEG preprocessing

All EC and EO resting- state EEG data, including locally acquired datasets (cohort 1 at all-time points, cohort 2 baseline data, and the healthy control short-term test–retest dataset) as well as the publicly available EEG dataset, were preprocessed using EEGLAB (v2024.2) ^50^ in MATLAB (R2022b) following a standardized pipeline.

For locally acquired datasets, electrodes not used for EEG signal analysis, specifically CPz, which was the reference electrode, along with M1 and M2, were removed, leaving 61 standard scalp channels for analysis (Fig. S2). Data were down-sampled to 250 Hz. 50-Hz Line noise was removed with a notch filter (49–51 Hz), which applied a zero-phase Hamming-windowed sinc FIR filter. A 1-Hz high-pass FIR filter was employed to eliminate slow drifts, while a 60-Hz low-pass FIR filter was utilized to suppress high-frequency noise.

Independent component analysis (ICA) was performed using the Infomax algorithm^49^. The number of ICA components was determined by eigenvalue decomposition of the data covariance matrix, retaining components until the cumulative eigenvalue sum reached 99% of total variance. Each independent component was automatically labeled by the ICLabel toolbox^51^, and components with an ocular (eye movement/blink) contribution greater than 0.7 were removed. Noisy time segments were identified using a sliding-window method (1 s window length) combined with an amplitude thresholding rule: if at least half of the data points in at least five channels exceeded 100 *µV* within a given window, that window was marked for rejection^52^.

Noisy channels were further identified using a robust procedure based on IQR. For each channel, we calculated the IQR of the band-pass–filtered amplitude distribution. To determine whether a channel should be rejected, its IQR was compared against a condition-specific threshold. These thresholds were derived from the pooled distribution of IQR values across all channels and participants within each condition (EO or EC). Specifically, the rejection threshold was defined as

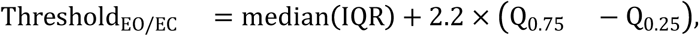

where Q_0.75_ and Q_0.25_ denote the first and third quartiles of the pooled IQR distribution, respectively. Channels exceeding this threshold were considered bad. Besides, spectral characteristics were also used to identify noisy channels. Specifically, the aperiodic exponent—the slope of the 1/f-like background activity in the neural power spectrum—was calculated, as it reflects the balance of excitation and inhibition in cortical circuits^48^. Channels with an aperiodic exponent > 3 were flagged as bad, since physiologically plausible values typically range from ∼0.5 to 3.0 in EEG/MEG recordings^48^. Datasets in which more than 30% of channels were flagged by either criterion were considered excessively noisy and excluded. For datasets not meeting this exclusion criterion, the identified bad channels were interpolated using spherical splines.

After artifact correction, data were re-referenced to the common average reference (CAR). Finally, a manual review was independently performed by two trained operators to remove residual bad channels and transient high-amplitude noise not captured by automated methods. Based on inspection of power spectra and EEG time-series traces, channels with abnormal frequency characteristics and time segments containing ocular artifacts or transient high-amplitude noise were identified. Only those channels and segments judged as artifactual by both operators were removed, ensuring that the procedure minimized non-physiological artifacts while preserving neural signals of interest.

The publicly available healthy-control EEG dataset was acquired using different hardware (a 64-channel cap with FCz as the reference). Consequently, only the initial channel selection step differed: all recorded channels were retained for analysis rather than excluding mastoid and reference electrodes as in the local datasets. All subsequent preprocessing steps—including resampling to 250 Hz, notch filtering, high- and low-pass filtering, ICA decomposition with ocular component removal, bad channel detection and interpolation, and common average referencing—were performed identically. Final manual review for residual artifacts and bad segments was also performed by the same trained operators, ensuring consistent artifact handling and quality control across both local and public datasets.

### Aperiodic exponent analysis

Aperiodic spectral activity was estimated from the preprocessed resting-state EEG data using FOOOF^48^. For each participant and condition, the cleaned EEG data were loaded from EEGLAB .set files, and power spectral density was calculated for each channel using Welch’s method. The spectra were restricted to the 1-50 Hz range before model fitting. FOOOF was then applied to each channel spectrum to parameterize the broadband aperiodic component and separate it from putative oscillatory peaks. The offset and aperiodic exponent were extracted for each channel, with the exponent used as the primary non-oscillatory spectral feature. Oscillatory peak frequency, power, and bandwidth were also retained when present. Finally, the channel-wise exponent values were organized separately for the eyes-open and eyes-closed conditions for subsequent analyses.

### MEG preprocessing

Resting- state MEG data were processed in MNE-Python using the same pipeline for both magnetometer (MAG) and gradiometer (GRAD) sensors. Data were down-sampled to 250 Hz, and notch (50 Hz), high-pass (1 Hz), and low-pass (60 Hz) filters were applied. ICA was used to separate noise components, with ECG and EOG signals serving as references for automatic artifact identification (correlation coefficient > 0.4).

### Structural MRI segmentation and co-registration

Because neural currents propagate differently through head tissues, structural MRI segmentation was required for MEG source analysis. FreeSurfer5 was used for automated segmentation of scalp, skull, cortex, and white matter. The pipeline included bias-field correction, skull stripping, intensity normalization, probabilistic tissue labeling, and cortical surface reconstruction. Segmentation outputs provided 3D models of scalp, skull, cortex, and white matter for individualized head modeling.

Segmented data were imported into Brainstorm, and anatomical landmarks (nasion, left and right preauricular points) and additional fiducials (anterior commissure, posterior commissure, inter-hemispheric point) were manually identified for structural alignment. To account for individual anatomical variability, structural images were further aligned to MNI space using SPM’s mutual-information-based affine registration.

### MEG source localization

Preprocessed resting- state MEG data were co-registered with each participant’s individual structural MRI and head geometry using digitized landmarks, followed by optimization based on the recorded head position. Individualized head models were then built using the overlapping-spheres method, which is a computationally efficient technique that locally approximates electromagnetic properties under each sensor. The resulting cortical source space consisted of ∼15,000 vertices, each modeled as a dipole constrained to be normal to the cortical surface.

Source estimation was performed using minimum norm estimation (MNE), combining sensor-level signals from both magnetometers and gradiometers and the lead-field matrix to estimate cortical current densities. The estimated source activity was then averaged within predefined regions of interest (ROIs), yielding ROI time series for computing log relative power features analogous to EEG. Cortical parcellation was based on the Destrieux atlas (148 gyral and sulcal regions)^37^.

### Normalized positive predictive value (nPPV) calculation

To quantify the relative improvement in predictive precision, nPPV^17^ were used, providing a measure of the proportional gain in predictive value attributable to biomarker stratification compared with the overall cohort, and defined as:

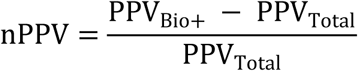

where PPV_Bio+_ is the positive predictive value within the biomarker-positive subgroup and PPV_Total_ is the overall cohort-level PPV.

### Log relative power feature extraction

For EEG, band power was computed for each channel and normalized to the broadband (1–50 Hz) power to obtain relative power. For example, for the delta band:

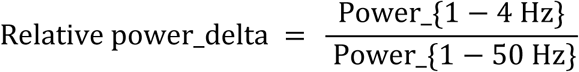

The resulting values were log-transformed to yield log relative power features:

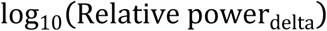

For MEG, source-space signals were derived using the above source reconstruction, averaged by ROI, and processed with the same log relative power feature extraction method.

### Nested leave-one-subject-out cross-validation (LOSOCV) procedure

To ensure an unbiased assessment of prediction performance, a nested cross-validation (LOSOCV) procedure was implemented to identify and assess the most predictive feature for symptom change, defined as the 6-month reduction rate in Y-BOCS scores.

The candidate feature set comprised log-transformed relative power values from five frequency bands—delta, theta, alpha, beta, and gamma—across 61 EEG channels and two resting-state conditions (EC and EO), yielding a total of 610 features. The nested LOSOCV procedure proceeded as follows:

1. **Outer Loop (Performance Assessment)**: In each iteration, one subject is held out as the final test subject, while the remaining 21 subjects form the training set.
2. **Inner Loop (Feature Selection)**: In each iteration of the outer loop, a separate LOSOCV is performed exclusively within the training set to identify the best feature. This inner loop evaluates each of the 610 candidate features by its ability to predict outcomes using the least-squares linear regression model, selecting the one that achieves the lowest mean squared error (MSE).
3. **Model Training and Prediction**: The single optimal feature identified by the inner loop is then used to retrain a least-squares linear regression model on the entire 21-subject training set. This model then makes a prediction for the one subject held out by the outer loop.

This entire procedure consists of 22 iterations, so each subject serves as the test subject exactly once. Finally, the overall performance of the biomarker is calculated by computing the Pearson’s correlation between the aggregated predictions and the true outcomes. This nested structure ensures that feature selection is performed strictly within the training data for each outer loop, thereby avoiding information leakage and producing an unbiased estimate of how well our biomarker would generalize to a new, unseen subject.

The predicted 6-month Y-BOCS reduction rates from the outer-loop test sets were correlated with the actual observed reductions using Pearson’s correlation coefficient. Statistical significance was assessed via permutation testing, with the *p*-value calculated as the proportion of permutations yielding a correlation equal to or greater than the observed value.

### Correlation and significance testing

Pearson correlation coefficients were calculated between frequency-band features and clinical outcome (6-month Y-BOCS reduction rate). Statistical significance was assessed using a non-parametric permutation test (5,000 permutations). Clinical labels were randomly permuted, correlations recomputed, and the resulting distribution of permuted statistics used as the empirical null. Empirical p-values were obtained as the proportion of permuted correlations equal to or more extreme than the observed statistic, thereby minimizing distributional assumption^53^.

### Test–retest reliability analysis

Test–retest reliability was assessed using the intraclass correlation coefficient [ICC(3,1); two-way mixed-effects model, single measurement, consistency definition]^38^. For a design with n subjects each measured at k sessions, the ICC(3,1) is defined as:

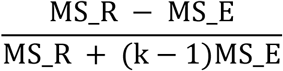

where MS_R is the mean square for rows (between-subject variance), and MS_E is the mean square error (within-subject variance). This formulation quantifies the proportion of total variance attributable to between-subject differences, reflecting the reliability of a single measurement across sessions. Values closer to 1 indicate excellent stability, while values near 0 suggest poor reproducibility. Analyses were conducted in three independent datasets:

1. **OCD discovery cohort.** Resting-state EEG in the EC condition was recorded preoperatively and again 1–3 months postoperatively, prior to DBS initiation. One patient did not undergo postoperative EEG acquisition before stimulation was initiated, resulting in 21 patients included in the reliability analysis.
2. **Healthy control dataset 1.** EEG data were obtained from a randomized controlled trial including a healthy control group (NCT06692114). 40 participants were enrolled, and 39 had complete data across two sessions separated by two weeks, acquired under identical laboratory conditions (same site, equipment, and personnel as the OCD cohort).
3. **Healthy control dataset 2.** A subset of 208 underwent repeat EEG recordings approximately five years after baseline, enabling assessment of long-term stability. EC recordings were obtained both before and after cognitive task performance at each session.

All datasets underwent identical automated preprocessing followed by manual quality control.

### Predictive modeling

For treatment prediction, a linear regression model was constructed using the preoperative EEG signature (i.e., right fronto-temporal delta relative power) as the predictor and 6-month Y-BOCS reduction rate as the dependent variable in the discovery cohort. The model was fitted using the least-squares approach and applied directly to independent validation subjects, requiring only preoperative EEG input, independent of clinical variables or manual judgment, ensuring interpretability and reproducibility. Model predictions were compared with observed clinical outcomes to assess generalizability.

### PLS-based transcriptomic association analysis

We first performed EEG source reconstruction to obtain cortical source activity estimates. The cortical surface was then parcellated into 148 regions of interest (ROIs) using the Destrieux atlas. For each ROI, we computed the mean baseline log-transformed delta-band relative power in source space. Across subjects, we then quantified the Pearson’s correlation between this ROI-level delta power and clinical outcome, defined as 6-month Y-BOCS reduction rate. This yielded one scalar per cortical region, reflecting how strongly that region’s baseline electrophysiological activity predicts subsequent clinical benefit. We refer to this regional map as the source-space EEG treatment-response correlation map. For downstream analyses, we specifically focused on the 30 ROIs exhibiting the strongest negative correlations with clinical outcome.

Regional cortical gene expression was obtained from the Allen Human Brain Atlas^54^ via the abagen toolbox^55^, which aggregates bulk microarray expression profiles sampled from the human cortex and is widely used to relate macroscale neuroimaging maps to underlying transcriptional architecture.

To move from a spatial EEG signature to a molecular mechanism, we next asked which cortical genes track the source-space EEG treatment-response correlation map. Before modeling, both the regional gene expression matrix (cortical ROIs * genes) and the source-space EEG treatment-response phenotype (one value per ROI) were z-scored across regions to place all variables on comparable scales and to remove arbitrary amplitude differences. We then performed partial least squares (PLS^56^) regression to relate regional cortical gene expression to the source-space EEG treatment-response phenotype. The first component (PLS1) was retained as the dominant axis because it explained the largest share of phenotype variance and remained significant under spatial permutation testing (permutation *p* < 0.05), and its regional PLS1 scores were strongly correlated with the source-space EEG treatment-response phenotype.

To link the PLS-derived cortical gene ranking to cellular mechanisms, we performed pre-ranked gene set enrichment analysis (GSEA) on the bootstrap-stabilized, sign-aligned PLS Z-scores (positive values indicate stronger association with greater clinical benefit in the EEG treatment-response phenotype). Destrieux ROIs were harmonized with AHBA parcels, ROIs without phenotype values were removed, genes with non-numeric/missing expression across parcels or zero across-parcel variance were excluded, and probe IDs were mapped to unique HGNC symbols with duplicates per symbol collapsed by retaining the entry with the largest |Z| in the PLS ranking. The resulting gene set defined the analysis universe. Gene-set libraries consisted of broad brain cell-type marker sets curated from CellMarker 2.0-excitatory neuron, inhibitory neuron, astrocyte, microglia, oligodendrocyte, oligodendrocyte precursor cell, endothelial, pericyte, and ependymal. Each set was intersected with the universe and required a minimum overlap of more than 3 genes.

For GSEA, we used the standard running-sum statistic on the signed Z-score vector (descending by Z) with 20,000 permutations of the ranked list to compute normalized enrichment scores (NES). NES > 0 indicates that markers concentrate near the top of the PLS ranking; NES < 0 indicates depletion toward the bottom. Nominal p-values were derived from the permutation null and Benjamini–Hochberg FDR was applied across all tested sets (significance at FDR < 0.05). For reporting, we visualized NES, −log10(FDR), and set size using bubble plots, and provided leading-edge genes for significant terms.

## Data Availability

The data are not publicly available due to patient privacy and institutional restrictions

## Author contributions

Conceptualization: Z Wang, W Wu

Methodology: W Wang, J Cheng, W Wu, Z Wang

Investigation: H Han, Q Fan, Z Wang, J Cheng, H Ruan, B Huang, T Xu, J Gao, X Zhang, X Wu

Clinical assessment: J Cheng, H Ruan, B Huang, T Xu, J Gao

Surgery: X Zhang, X Wu

Data curation: W Wang, J Cheng, Z Wang

Formal analysis: W Wang, J Cheng

Validation: W Wu, Z Wang

Visualization: W Wang, J Cheng, Y Liang

Software: F Qi, H Ruan, H Zhi, Y Liang, B Huang, L Cao

Funding acquisition: Z Wang, W Wu

Supervision: W Wu, Z Wang

Project administration: W Wu, Z Wang

Writing – original draft: W Wang, J Cheng, W Wu

Writing – review & editing: W Wang, J Cheng, W Wu, Z Wang, H Ruan, C Keller, G Schalk, N Williams

## Acknowledgments

We are thankful to the participants of this study for their time and participation.

## Data and materials availability

Healthy control dataset 2 is publicly available via Openneuro (accession number: ds005385).

## Code availability

Code used is available from the authors upon reasonable request.

## Supplementary Materials list

Figs. S1 to S10

Tables S1 and S2

Supplementary Text

References (supplementary references 1 to 11)

