## Supplementary files for "A Preoperative Electroencephalography Signature for Predicting Treatment Response to Deep Brain Stimulation in Obsessive-Compulsive Disorder"

**Supplementary Materials**

This PDF file includes:

**Figs. S1 to S10**

**Tables S1 and S2**

**Supplementary Text**

Safety

Reduced delta power in Bio+ OCD patients versus healthy controls

Topographic analyses of relative power

Image acquisition and DBS lead localization

**Supplementary References (1 to 11)**

### Supplemental figures

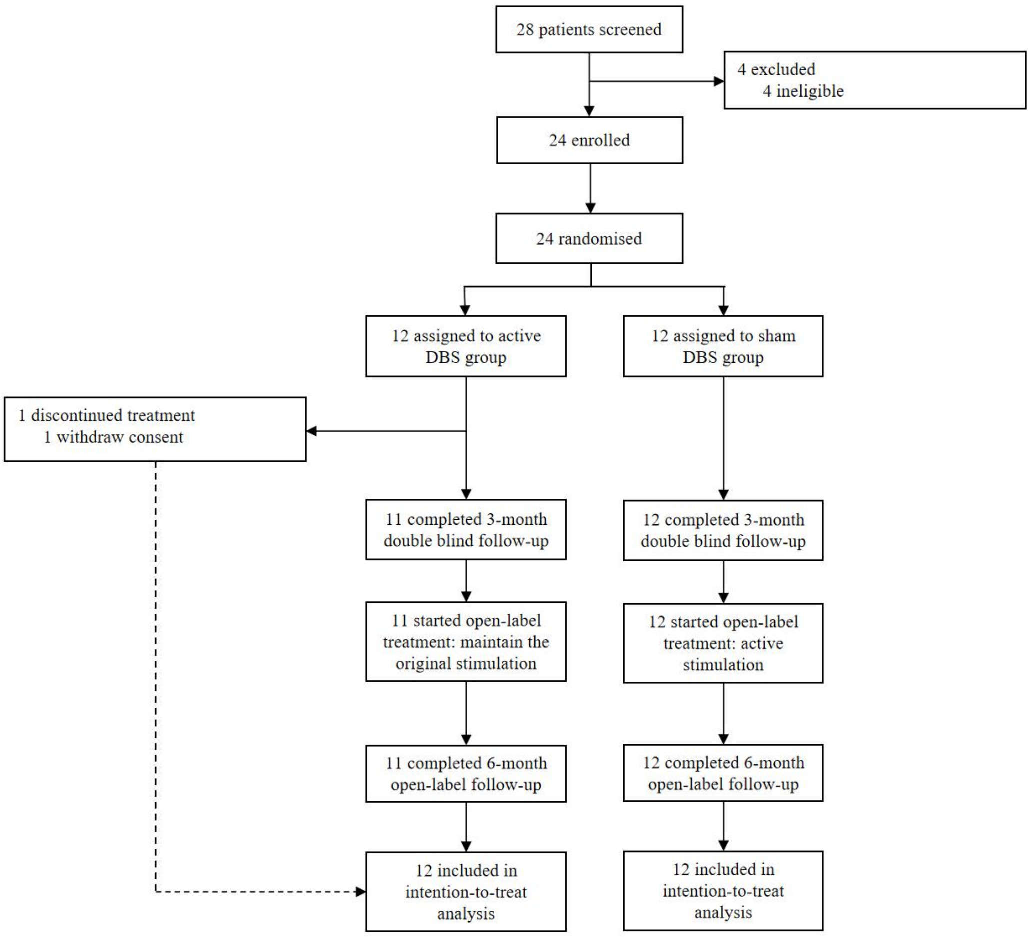


**Fig. S1.** CONSORT flow diagram of patient enrollment, randomization, and follow-up in the OCD DBS clinical trial.

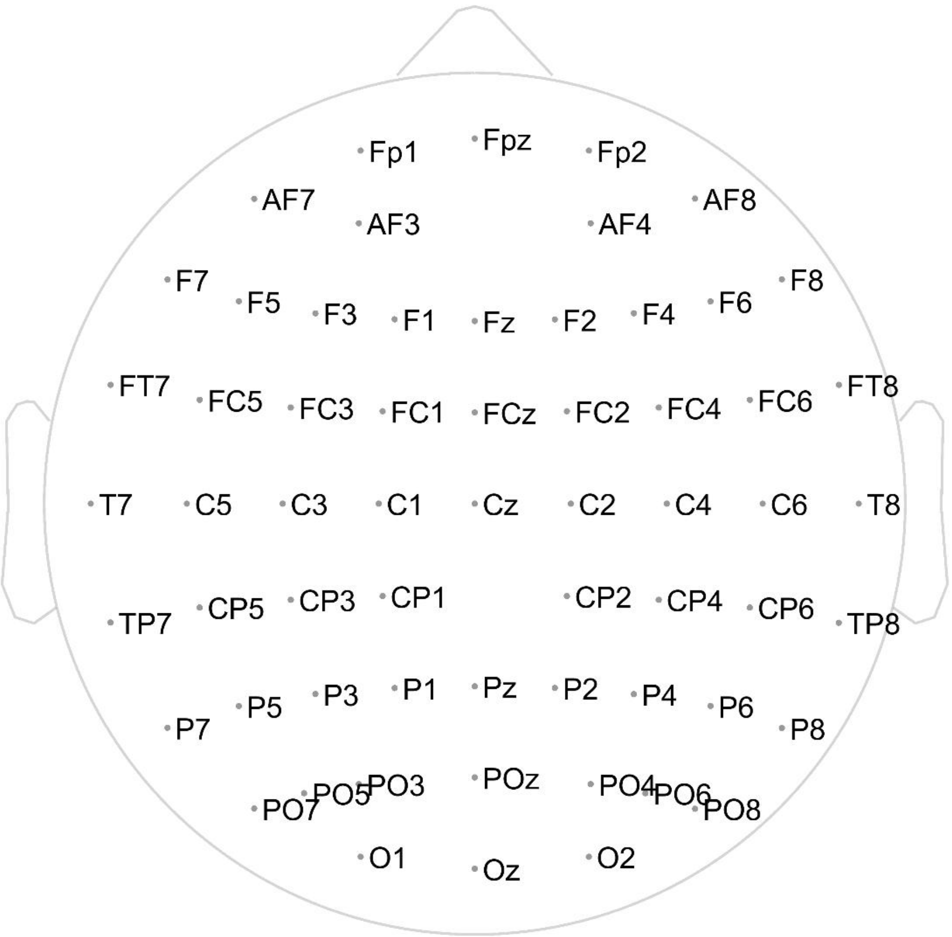


**Fig. S2.** Electrode layout used in the analysis of the dataset from our center.

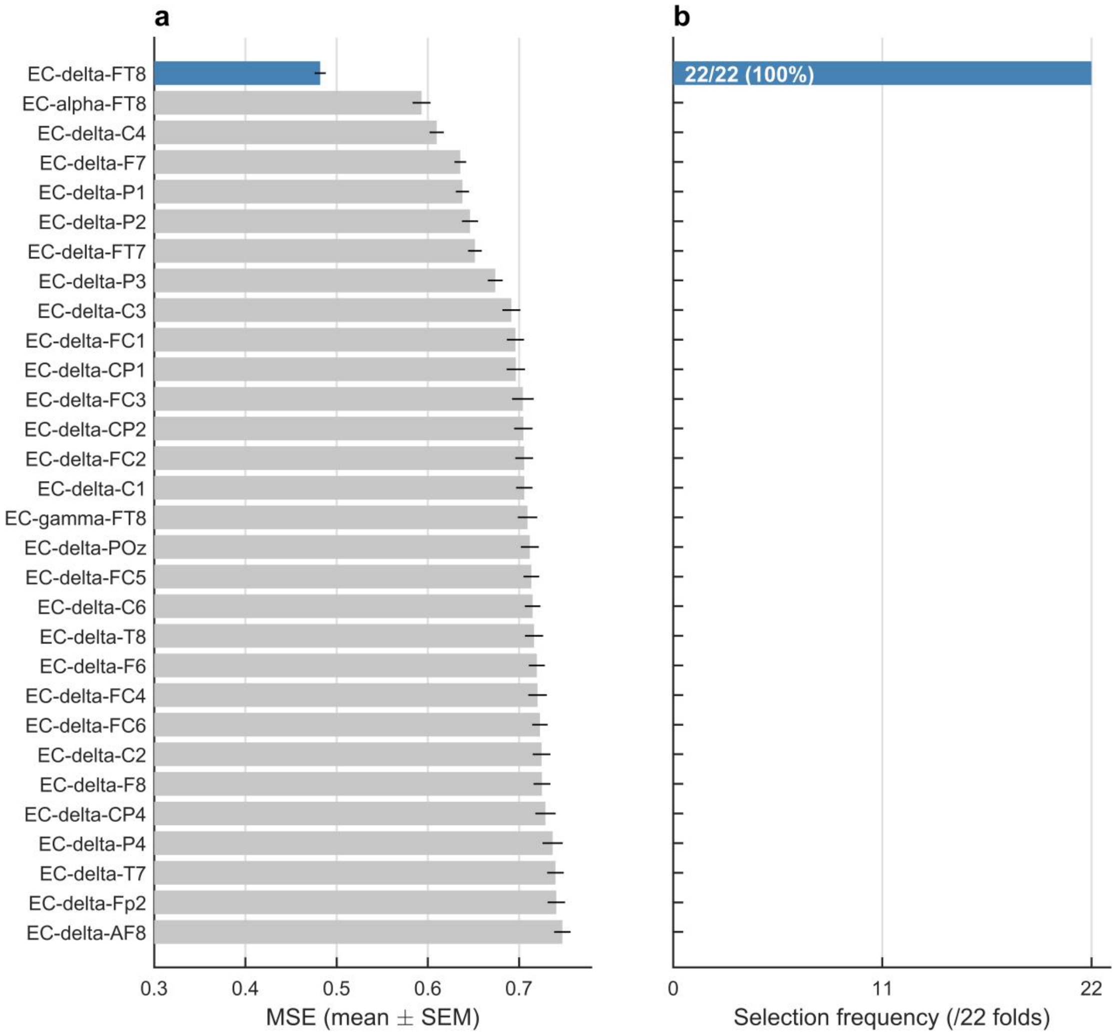


**Fig. S3. Cross-validated feature ranking and selection frequency in the nested LOSOCV. a**. Mean ± SEM cross-validated error (MSE) for the 30 best of 610 candidate features across the 22 outer folds; EC eyes-closed fronto-temporal (FT8) delta power (blue) had the lowest error. **b**. Number of outer folds (of 22) in which each feature was selected as the single optimal predictor. EC FT8 delta power was selected in all 22/22 folds.

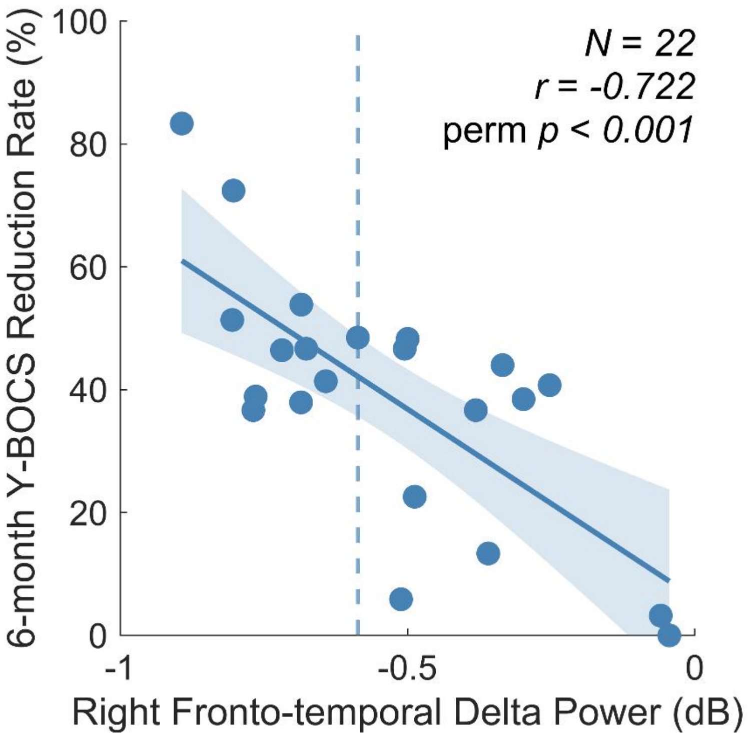


**Fig. S4.** Association between preoperative right fronto-temporal delta relative power and six-month Y-BOCS reduction rate. Each dot represents one patient ($N$ = 22). The dashed blue line indicates the biomarker-derived cutoff, while the solid blue line represents the regression fit with shaded area denoting the 95% confidence interval.

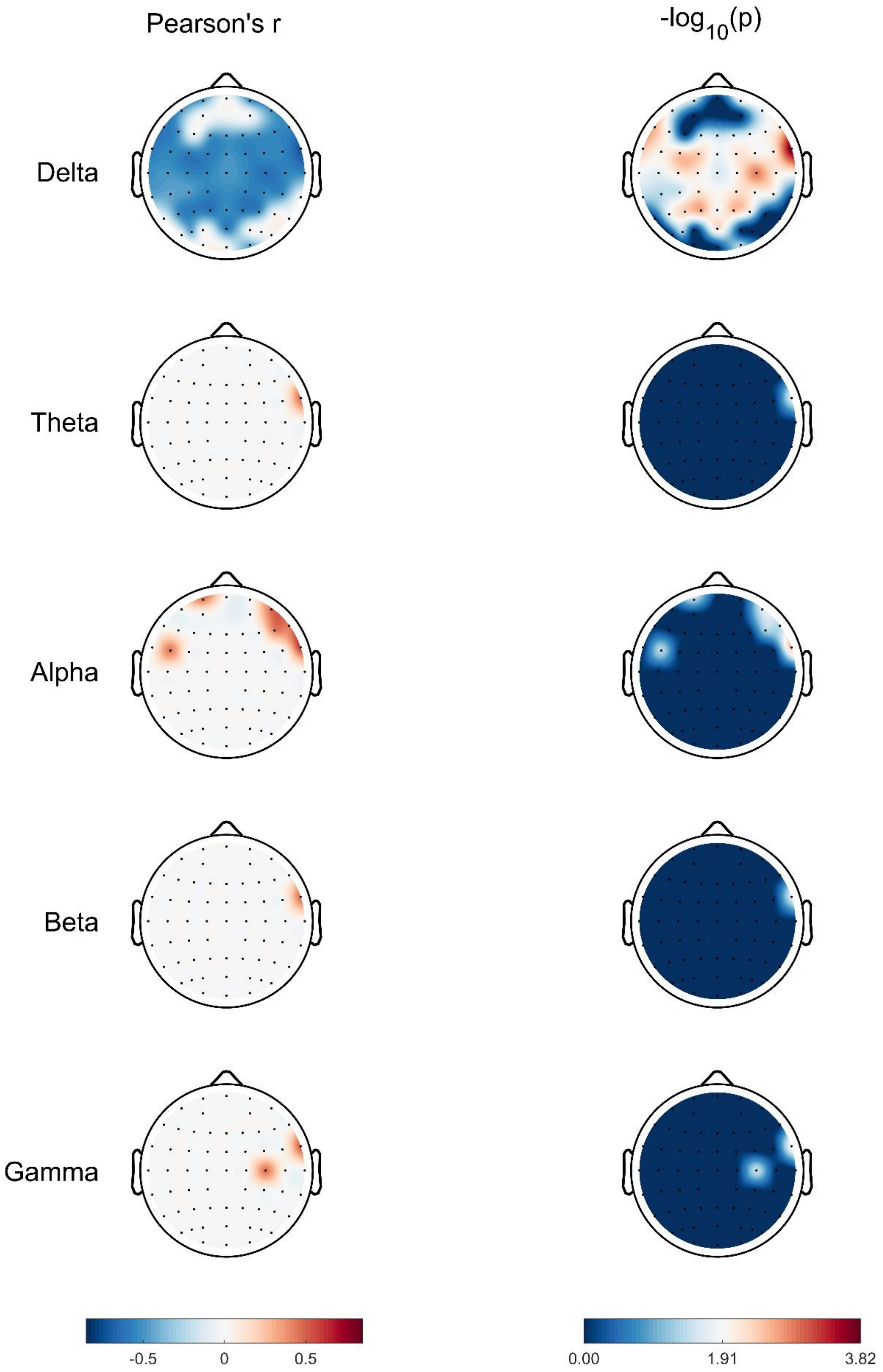


**Fig. S5.** Topographic distribution of Pearson correlation coefficients (Pearson's *r*) and statistical significance (-$\log_{10}$(permutation *p*)) across five frequency bands (delta, theta, alpha, beta, gamma) in the patient cohort ($N$ = 22). Only significant channels are displayed (permutation $p$ < 0.05).

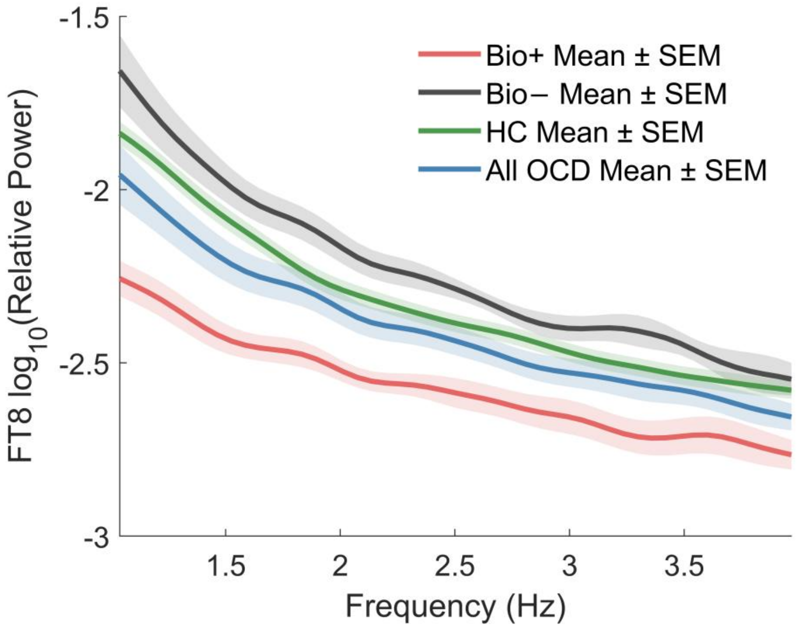


**Fig. S6.** EC FT8 delta-band relative power spectra. Group-level spectra (mean ± SEM) in biomarker-positive (Bio+, *N* = 11), biomarker-negative (Bio–, *N* = 11), all OCD patients (*N* = 22), and healthy controls (HC, *N* = 40).

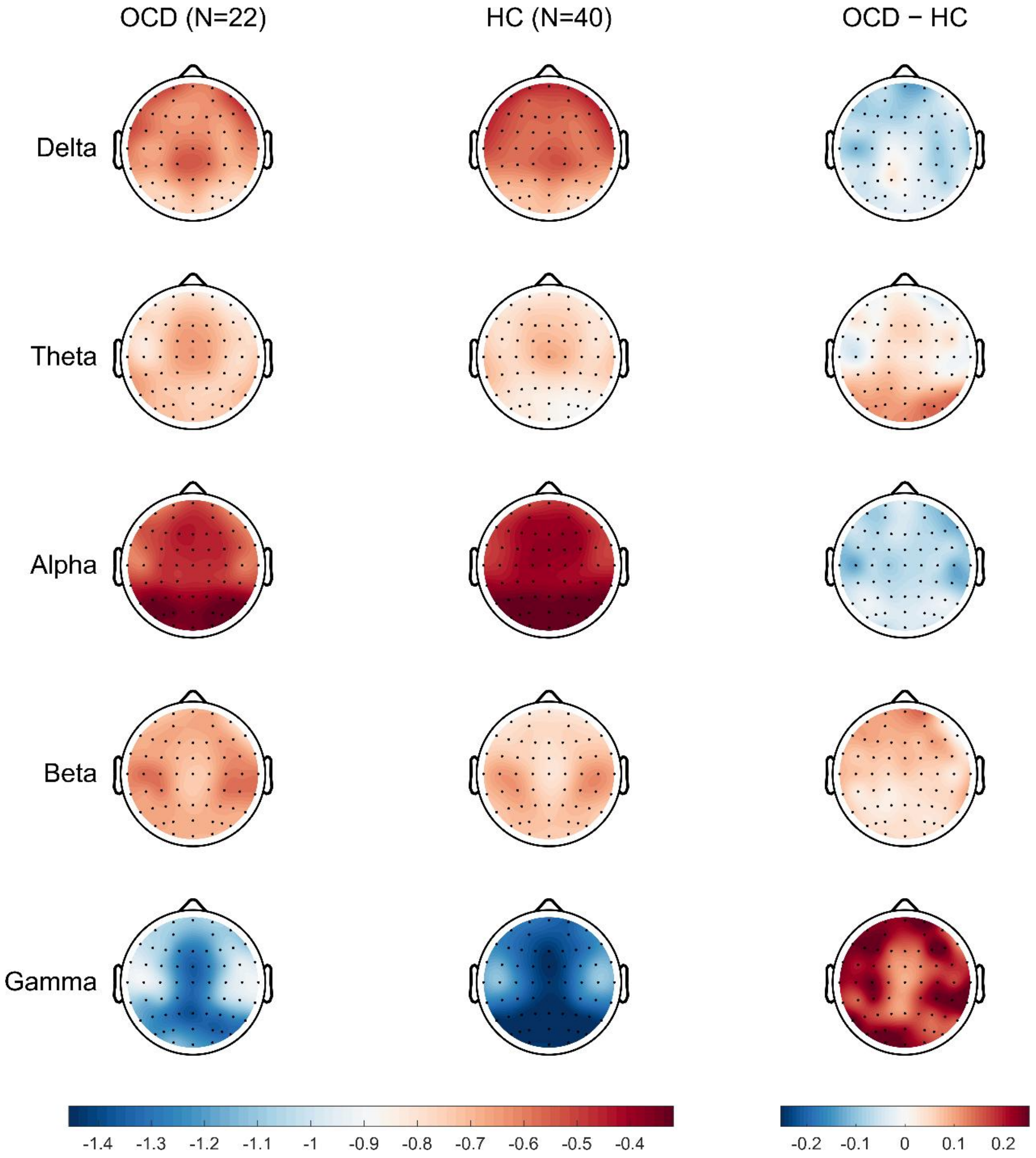


**Fig. S7.** Topographical maps of log-transformed relative power in OCD patients and healthy controls. OCD patients (left) and healthy controls (middle) show broadly similar spatial distributions across frequency bands, while subtraction maps (OCD – HC; right) highlight reduced delta and alpha power in OCD over prefrontal and right temporal regions, and increased gamma power relative to HC. Theta and beta band differences were modest and spatially variable.

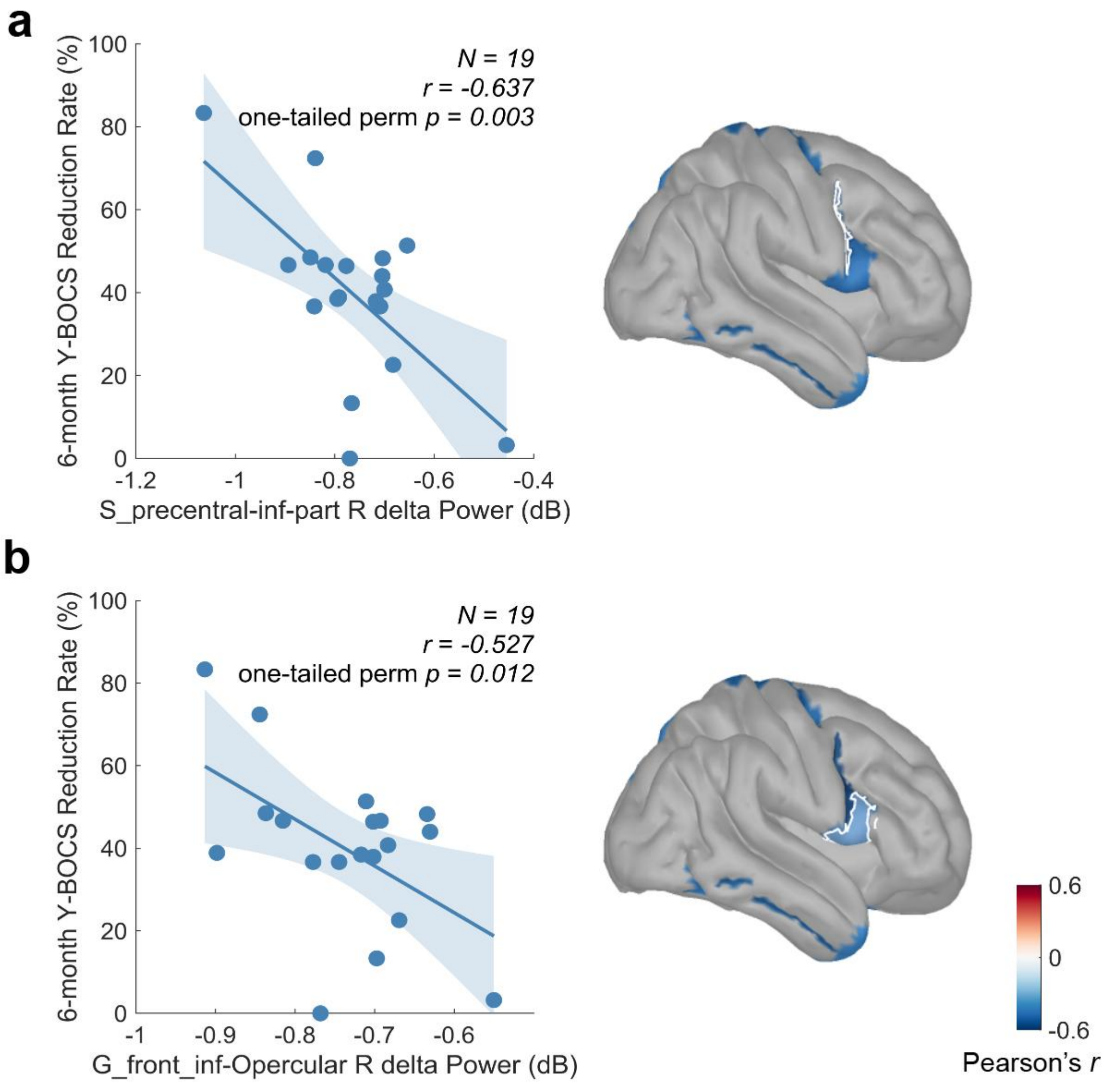


**Fig. S8.** Source-level MEG correlations in two significant right-hemisphere ROIs. **a**, Right inferior part of the precentral sulcus (S_precentral-inf-part R). **b**, Right opercular part of the inferior frontal gyrus (G_front_inf-Opercular R).

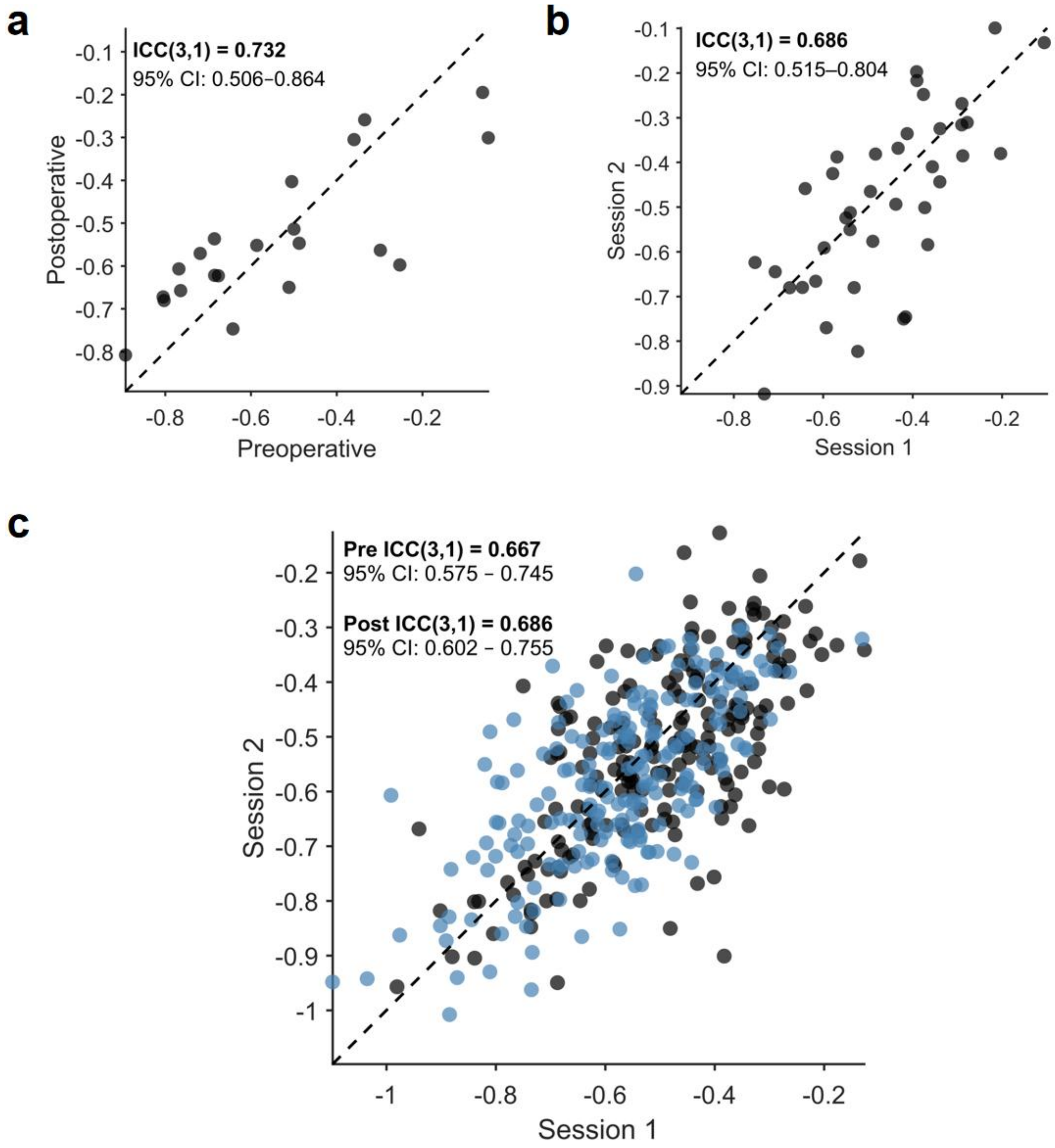


**Fig. S9.** Test–retest reliability of the EEG signature in OCD patients and healthy controls. a, Scatter plot of eyes-closed right fronto-temporal delta power measured preoperatively and postoperatively in patients in cohort 1 (*N* = 21). Each dot represents an individual patient; the dashed line indicates the line of identity. ICC values and 95% bootstrap confidence intervals are shown. b, Illustration of the two-week reliability assessment in healthy controls (*N* = 39). c, Long-term reliability in HC (*N* = 208) with repeat recordings at Session 1 and Session 2 (~5 years apart), shown separately for pre-task (blue) and post-task (dark) eyes-closed conditions.

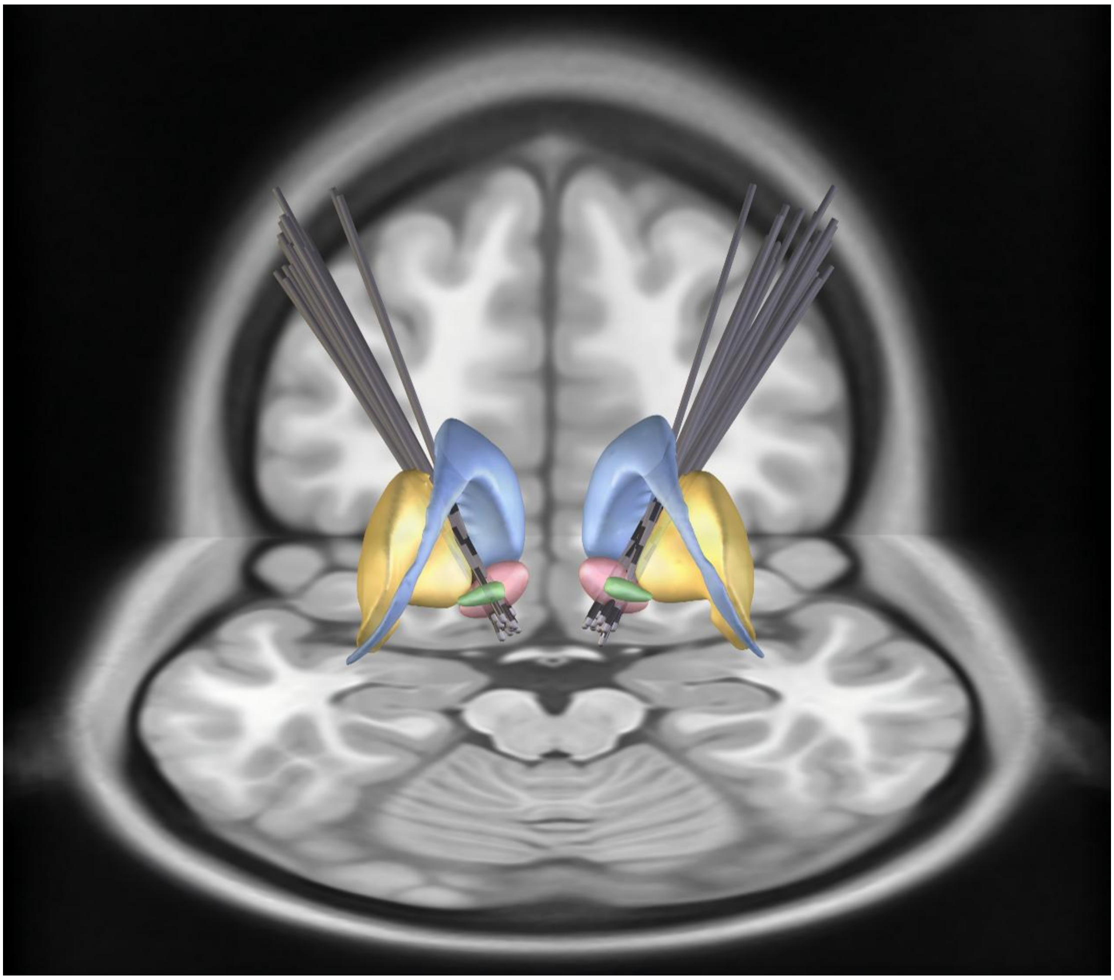

**Fig. S10.** Overview of lead electrode placement. The same patients from the discovery cohort (*N* = 22) are shown, with electrode contacts targeting the nucleus accumbens (NAcc) and the anterior limb of the internal capsule (ALIC). In the figure, red indicates the NAcc, green the ventral pallidum, yellow the putamen, and blue the caudate. Subcortical structures were defined by the CIT-168 Reinforcement Learning Atlas[1] (ALIC/NAcc region), with coronal and axial planes of the T1-weighted MNI152 2009b nonlinear template[2] as background.

**Supplemental Materials**

### Safety

Adverse events were described in table S2. During the RCT phase, a total of 31 adverse events (AEs) were reported in the active DBS group, compared with 17 AEs in the sham stimulation group. In the active DBS group, 10 AEs were device-related, and 10 device-related AEs were reported in the sham stimulation group. No serious adverse events (SAEs) were reported in either group. Only one participant in the active DBS group withdrew from the study before completion of the randomized phase due to withdrawal of informed consent. No patients withdrew due to AEs or SAEs.

### Bio+ OCD patient exhibit significantly reduced delta power relative to healthy controls

At the channel level, FT8 delta-band relative power spectra revealed that biomarker-positive (Bio+) patients exhibited significantly reduced delta power compared with healthy controls (HC), whereas biomarker-negative (Bio–) patients showed values comparable to HC (Fig. S4).

Topographic analyses of log-transformed relative power further demonstrated that the overall spatial distributions were broadly similar across frequency bands. However, subtraction maps (OCD – HC) revealed frequency-dependent alterations, including reduced delta and alpha power over prefrontal and right temporal regions, increased gamma power over frontal and parietal regions, and modest, spatially variable differences in theta and beta bands (Fig. S5). Importantly, these effects were primarily driven by the Bio+ subgroup, while the Bio– subgroup remained largely indistinguishable from HC.

### Image acquisition and DBS lead localization

For cohort 1 (*N* = 22), high-resolution structural T1-weighted MRI were acquired preoperatively on a 3T Siemens Verio scanner (3D MPRAGE; voxel size=1*1*1mm$^3$, repetition time (TR) = 2300 ms, time echo (TE) =3.5 ms, flip angle (FA) = 9°, field of view (FoV) = 256*256 mm$^2$). Postoperative computed tomography (CT) was obtained to verify electrode placement.

DBS electrodes were localized using the advanced processing pipeline[3,4] in Lead-DBS software (version 3.2; http://www.lead-dbs.org)[5]. Post-operative CT scans were linearly co-registered to preoperative T1 images using Advanced Normalization Tools (ANTs, http://stnava.github.io/ANTs/)[6]; when this registration failed, the FSL FLIRT[7] preset in Lead-DBS was used. Subcortical refinement was applied (as a module in Lead-DBS) to correct for brain shift that may have occurred during surgery. Images were then normalized to the ICBM 2009b Nonlinear Asymmetric (MNI) template[2] with the ANTs SyN algorithm[8]. A subcortical refinement step (Lead-DBS "Effective: Low Variance" preset) was applied to obtain a most precise subcortical alignment between patient and template space. Co-registrations and normalizations were visually inspected and refined as needed; where clear mismatches were present, deformation fields were manually adjusted with the WarpDrive[9] tool in Lead-DBS, with particular attention to the NAcc. Electrodes were localized in Lead-DBS and warped to MNI space; contacts were reconstructed with the automated PaCER[10] method and manually refined when necessary. Subcortical structures labels were taken from the CIT168 Reinforcement Learning atlas[1], and group visualizations were generated with Lead-Group[11].

**Table S1. Pearson correlation between baseline demographic and clinical variables and 6-month Y-BOCS reduction rate for cohort 1.**

| Predictor | *N* | *r* | *p*_perm | slope | intercept |
|---|---|---|---|---|---|
| Education | 22 | 0.215 | 0.336 | 0.015 | -0.720 |
| Sex | 22 | -0.118 | 0.600 | -0.066 | -0.454 |
| Age | 22 | -0.114 | 0.613 | -0.002 | -0.463 |
| BMI | 22 | 0.067 | 0.767 | 0.005 | -0.657 |
| Onset_age | 22 | -0.085 | 0.708 | -0.002 | -0.483 |
| Illness_duration | 21 | -0.155 | 0.504 | -0.005 | -0.467 |
| Y-BOCS | 22 | -0.141 | 0.533 | -0.011 | -0.208 |
| Obsession | 22 | -0.186 | 0.406 | -0.031 | -0.070 |
| Compulsion | 22 | -0.089 | 0.695 | -0.011 | -0.365 |
| CGI-S | 22 | -0.062 | 0.784 | -0.020 | -0.411 |
| HAMD | 22 | -0.083 | 0.712 | -0.003 | -0.494 |
| HAMA | 22 | 0.073 | 0.748 | 0.003 | -0.562 |

Y-BOCS = Yale-Brown Obsessive-Compulsive Scale, CGI-S = Clinical Global Impression -Severity, HAMD = Hamilton Depression Scale, HAMA = Hamilton Anxiety Scale

**Table S2. Adverse Events for cohort 1**

| | RCT phase | | Open-label phase (*N* = 23) |
|---|---|---|---|
| | Active DBS Group (*N* = 12) | Sham DBS Group (*N* = 12) | |
| SAEs | 0 | 0 | 0 |
| SAEs related to study device | 0 | 0 | 0 |
| SAE leading to discontinuation | 0 | 0 | 0 |
| Participants with any AE | 10 | 10 | 4 |
| Participants with any AE related to study device | 5 | 3 | 4 |
| Total AEs reported | 31 | 17 | 7 |
| AEs related to study device | 10 | 4 | 5 |

DBS = deep brain stimulation, RCT = randomized controlled trial, SAE: severe adverse event, AE = adverse event.
